# Occupational risks of COVID-19 in NHS workers in England

**DOI:** 10.1101/2021.04.08.21255099

**Authors:** DA van der Plaat, I Madan, D Coggon, M van Tongeren, R Edge, R Muiry, V Parsons, P Cullinan

## Abstract

**Objective:** To quantify occupational risks of Covid-19 among healthcare staff during the first wave of the pandemic in England

**Methods:** Using pseudonymised data on 902,813 individuals continuously employed by 191 National Health Service trusts during 1.1.19 to 31.7.20, we explored demographic and occupational risk factors for sickness absence ascribed to Covid-19 during 9.3.20 to 31.7.20 (n = 92,880). We estimated odds ratios (ORs) by multivariable logistic regression.

**Results:** With adjustment for employing trust, demographic characteristics, and previous frequency of sickness absence, risk relative to administrative/clerical occupations was highest in additional clinical services (including care assistants) (OR 2.31 [2.25-2.37]), registered nursing and midwifery professionals (OR 2.28 [2.23-2.34]) and allied health professionals (OR 1.94 [1.88-2.01]), and intermediate in doctors and dentists (OR 1.55 [1.50-1.61]). Differences in risk were higher after the employing trust had started to care for documented Covid-19 patients, and were reduced, but not eliminated, following additional adjustment for exposure to infected patients or materials, assessed by a job-exposure matrix. For prolonged Covid-19 sickness absence (episodes lasting >14 days), the variation in risk by staff group was somewhat greater.

**Conclusions:** After allowance for possible bias and confounding by non-occupational exposures, we estimated that relative risks for Covid-19 among most patient-facing occupations were between 1.5 and 2.5. The highest risks were in those working in additional clinical services, nursing and midwifery and in allied health professions. Better protective measures for these staff groups should be a priority. Covid-19 may meet criteria for compensation as an occupational disease in some healthcare occupations.

**Key messages:**

1. What is already known about this subject? *Healthcare workers and other keyworkers (workers whose job was considered essential to societal functioning) had a higher likelihood of testing positive for COVID-19 than other workers during the first lockdown in England. Amongst healthcare workers, those working in inpatient settings had the highest rate of infection*.
2. What are the new findings? *Between March and July 2000, the overall risk of COVID-19 sickness absence in National Health Service staff in England was lower at older ages, higher in non-white staff, and (in comparison with administrative and clerical staff) more than doubled in registered nurses and among workers such as healthcare assistants providing support to health professionals. Risk in health care scientists was little different from that in administrative and clerical occupations*
3. How might this impact on policy or clinical practice in the foreseeable future? *Our results suggest that the risk reduction strategies that were in place for healthcare scientists were effective. However, the protection for nursing and supporting health professionals was insufficient. In the event of a further ‘wave’ of infections resulting in high hospital admissions, attention should be paid to ensuring that risk reduction strategies for nurses and supporting health professionals are improved*.

## Introduction

Covid-19, like many communicable diseases, poses an occupational hazard to healthcare workers. When the first wave of the pandemic hit the UK early in March 2020, precautions were implemented to reduce transmission to healthcare staff, including identification and segregation of infected patients, and the use of personal protective equipment (PPE). In the early weeks, however, these measures were far from ideal. Adequate PPE often was in short supply and much of the UK’s pandemic stockpile contained equipment suitable for an influenza outbreak, but not for more infectious diseases (1). Additionally, a lack of capacity meant that testing of patients who might be carrying SARS-CoV-2 was insufficient (2). Cases of occupationally-acquired disease were therefore to be expected. However, the level of risk has been uncertain, as has the extent to which it varied between different healthcare occupations. Better understanding would help in prioritisation of preventive strategies during further waves of the pandemic, and in the management of similar infectious diseases. It is also needed to inform decisions on possible compensation for Covid-19 as an occupational disease in healthcare workers.

Evidence to date has indicated that male healthcare workers (taken as a group), nurses, nursing assistants and auxiliaries of both sexes have had higher age-adjusted mortality from Covid-19 in England and Wales compared with the general population (3). Several studies have found that patient-facing healthcare workers were infected with COVID-19 at substantially higher rates than non-healthcare workers during the first wave (March–July 2020), although studies differ in their findings as to which groups had been at greatest risk (4-6). Mortality, however, depends not only on risk of contracting Covid-19, but also on personal vulnerability when infection occurs, which may vary importantly between occupations. Furthermore, differences in the incidence of infection by occupation may be driven not only by exposures in the workplace (through proximity to infected colleagues as well as contact with patients and infected materials), but also away from work. For example, rates of infection have been higher among people living in large, crowded households (7)

To get further insight regarding occupational risks of Covid-19 in healthcare workers, we analysed data on sickness absence among employees of National Health Service (NHS) trusts in England, before and after they started to care for patients known to have the disease.

## Methods

With approval by the NHS Health Research Authority (reference 20/SC/0282), we were allowed access to two pseudonymised databases prepared by the NHS Electronic Staff Record (ESR) Central Team. They contained information on demographic and occupational characteristics of all staff continuously employed by NHS trusts in England from 01.01.2019 to 31.07.2020, and on all their absences from work during that period, other than for annual leave. The latter included the reason for absence, and the start and end date of each episode.

Supplementary File A describes the methods by which we used the two databases to create a file for statistical analysis. We first checked for missing and inconsistent data, and corrected clear anomalies in a small minority of records by imputation according to a standard set of rules. We also reclassified some variables into aggregated categories that would facilitate more meaningful analysis. We then generated a file with one record for each individual, which included the variables listed in Table 1, and also the start and end dates of all absences during 01.01.2019 to 31.07.2020, with the reason for absence.

**Table 1.**
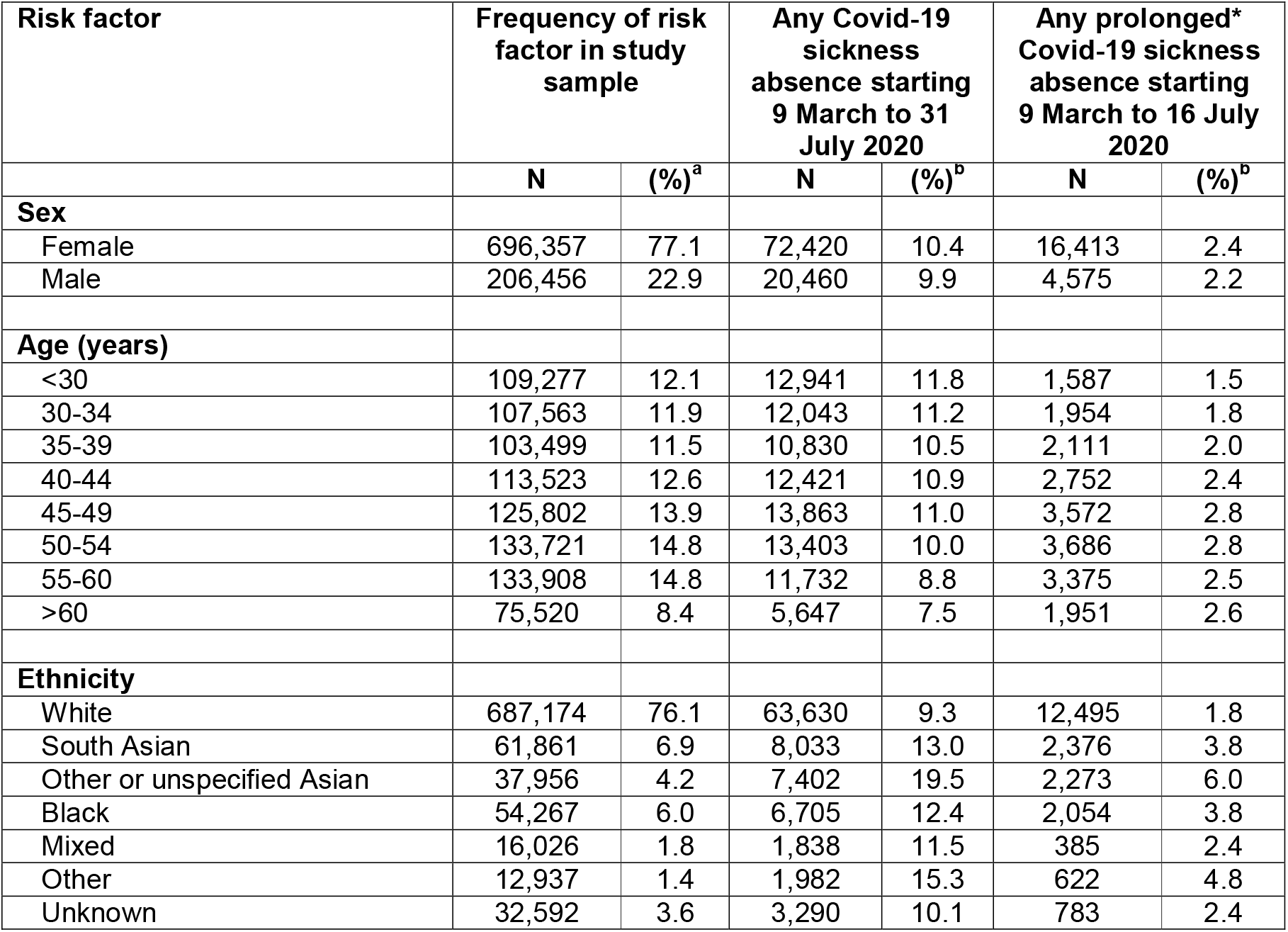
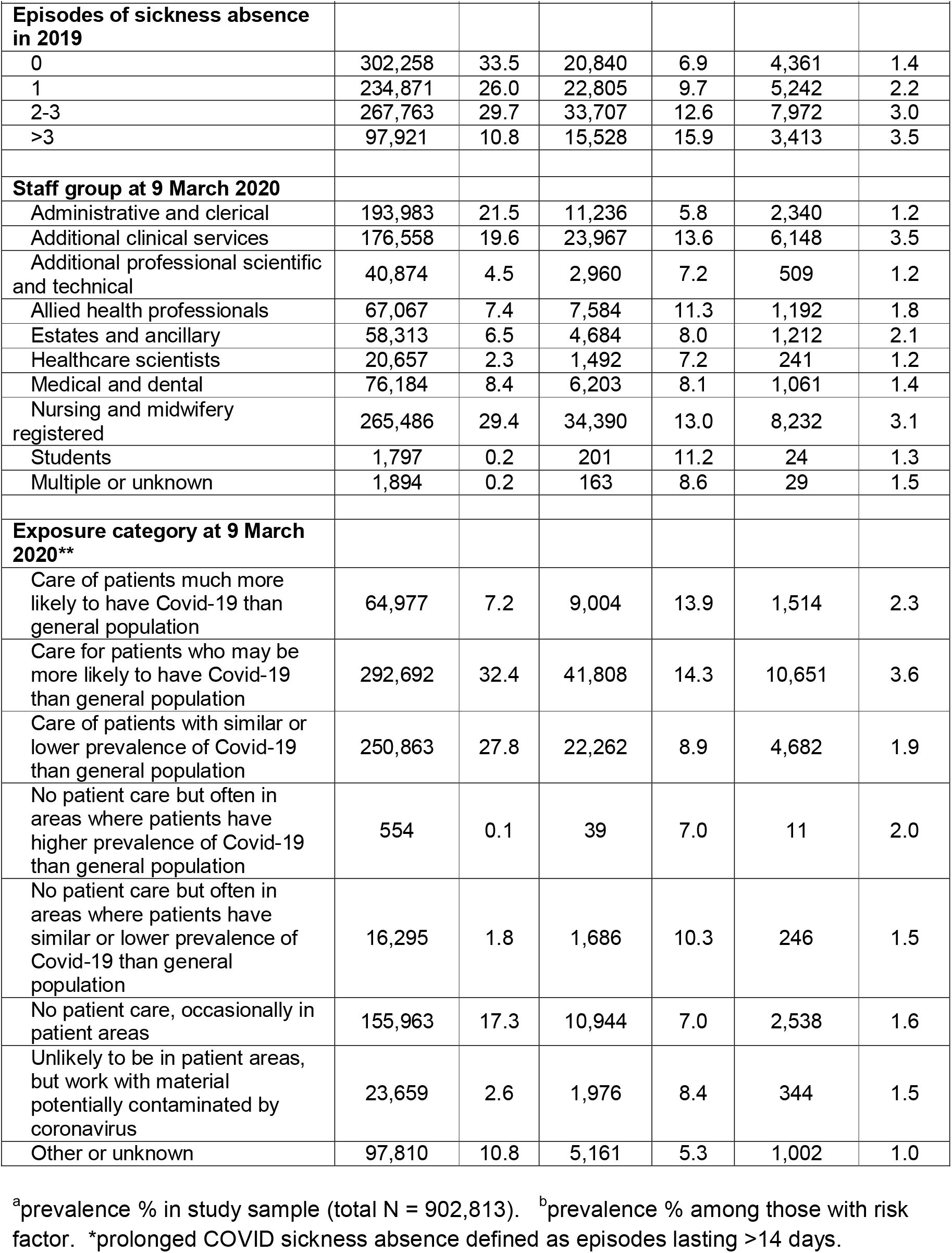
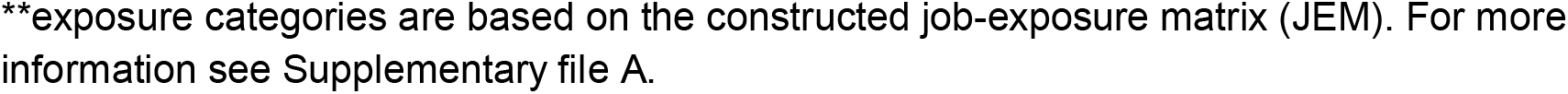
Distribution of risk factors in study sample and cumulative prevalence of new Covid-19 sickness absence during 9 March to 31 July 2020.

Staff group was assigned to 10 categories, according to a classification used in the ESR records (Supplementary File A – Table A1). The ESR system also held more detailed occupational data but to protect privacy that could not be released. Instead, four members of the team (an occupational hygienist and three occupational physicians with experience in the NHS) compiled a job-exposure matrix (JEM), which the ESR Management Team then used to reclassify detailed occupational categories (n=659) to the eight exposure categories listed in Table 1.

Within the ESR database, the reason for any type of absence was described by four variables (Supplementary File A). The 192 different combinations were collapsed into 60 categories, of which 32 were related to sickness absence. Using the information on absence episodes, we defined a variable which for each individual represented the number of new episodes of sickness absence (for any cause) that had started during 2019 (classified as 0, 1, 2-3 and >3). This was intended as a marker for long-term propensity to take sickness absence, which can vary importantly between individuals independently of morbidity (8). In addition, we distinguished episodes of Covid-19 sickness absence, which we defined as being for any of five categories of sickness (cough/flu, chest/respiratory, infectious diseases, other or unknown) with Covid-19 recorded as a related reason. Such episodes were classed as prolonged if their duration exceeded 14 days (see Supplementary File B).

Data on the date by which each trust was known to have admitted at least three Covid-19 cases were obtained from an NHS COVID-19 daily situation report published on 12.11.2020 (9). We took 09.03.2020 as the date from which Covid-19 sickness absence could reasonably be assumed to reflect coronavirus infection. That was at least 10 days before most hospitals started to admit documented Covid-19 cases (see Supplementary File B for further justification).

Two collaborating trusts provided data on antibody tests that had been carried out on staff members before 07.08.2020. Individuals were identified by an encrypted code number that had been assigned by the ESR Management Team, allowing anonymised linkage with the other records to which we had access.

### Statistical analysis

Statistical analysis was carried out with R (version 4.0.4) software. We first generated descriptive statistics summarising the distributions of the main variables. We then fitted two multivariable logistic regression models to estimate odds ratios (ORs) with 95% confidence intervals (95%CIs) for the start of any episode of Covid-19 sickness absence from 09.03.2020 to 31.07.2020. Model 1 included sex, age group, ethnicity, episodes of sickness absence in 2019 and staff group, while in model 2 the exposure category variable was additionally included to help understand the extent to which associations with staff group reflected patient-related exposures.

Next, the analysis was repeated, distinguishing between onset of the Covid-19 sickness absence before and after the employing trust had first cared for at least three documented Covid-19 cases. Our aim was to distinguish periods when acquisition of Covid-19 through transmission from patients was less and more likely; we incorporated a lag of four days to allow for an interval between exposure to infection and development of symptoms.

Further logistic regression models were used to explore risk factors for prolonged Covid-19 sickness absence starting during 09.03.2020 16.07.2020 (because records were complete only up to 31.07.2020, we could not be confident of accurately distinguishing prolonged episodes that started after 16.07.2020).

Finally, to check on the reliability of Covid-19 sickness absence as a marker for the disease, we used data from two collaborating trusts to compare the prevalence of positive antibody tests in employees who underwent testing before 07.08.2020, according to their history of Covid-19 sickness absence.

As sensitivity analyses, we excluded individuals in whom one or more of the age, sex or ethnicity variables was imputed because of inconsistencies, individuals with multiple jobs or whose job changed over the study period, or individuals with missing or imputed end date of an absence.

## Results

After exclusion of 21,775 employees who were absent from work continuously from 09.03.2020 to 31.07.2020 (mainly because of maternity or study leave), and 56,543 at nine trusts which never coded whether sickness absence was related to Covid-19, analysis was based on 902,813 individuals (77% female) from 191 trusts. Most (89%) were aged between 25 and 60 years, and 76% were of white ethnicity. A total of 92,880 (10%) had one or more episodes of Covid-19 sickness absence during the study period, including 20,988 (2.3%) in whom at least one episode was prolonged. Table 1 gives further information about the distribution of risk factors in the study sample, and the cumulative prevalence of Covid-19 sickness absence over the study period, according to those risk factors.

Table 2 shows associations of Covid-19 sickness absence at any time during the study period with the main risk factors of interest. After adjustment for other covariates, risk was similar in men and women, and in age groups below 55 years, but lower at older ages (OR for age >60 relative to <30 years in fully adjusted model: 0.76). Risk was generally higher for non-white relative to white ethnicity, and particularly for those of Asian origin (ORs 1.43 and 1.73 in fully adjusted model). Frequency of sickness absence during 2019 was a further risk factor, with an OR of 2.41 for >3 relative to 0 episodes in the fully adjusted model.

**Table 2.** Associations of risk factors at baseline with start of any Covid-19 sickness absence during 9 March to 31 July 2020. Risk estimates were derived from two logistic regression models that included all of the variables for which results are presented, together with trust (191 categories).

| Risk factor | Model 1 |  | Model 2 |  |
| --- | --- | --- | --- | --- |
|  | OR | (95% CI) | OR | (95% CI) |
| **Sex** |  |  |  |  |
| Female | *ref.* | *ref.* | *ref.* | *ref.* |
| Male | 1.01 | 0.99 - 1.03 | 1.02 | 1 - 1.03 |
| **Age (years)** |  |  |  |  |
| <30 | *ref.* | *ref.* | *ref.* | *ref.* |
| 30-34 | 0.96 | 0.94 - 0.99 | 0.97 | 0.94 - 0.99 |
| 35-39 | 0.97 | 0.95 - 1.00 | 0.98 | 0.96 - 1.01 |
| 40-44 | 0.99 | 0.96 - 1.02 | 1.00 | 0.97 - 1.02 |
| 45-49 | 1.00 | 0.98 - 1.03 | 1.00 | 0.98 - 1.03 |
| 50-54 | 0.98 | 0.95 - 1.00 | 0.98 | 0.95 - 1.00 |
| 55-60 | 0.89 | 0.86 - 0.91 | 0.89 | 0.86 - 0.91 |
| >60 | 0.76 | 0.74 - 0.79 | 0.76 | 0.73 - 0.79 |
| **Ethnicity** |  |  |  |  |
| White | *ref.* | *ref.* | *ref.* | *ref.* |
| South Asian | 1.43 | 1.4 - 1.47 | 1.41 | 1.37 - 1.45 |
|  | OR | (95% CI) | OR | (95% CI) |
| Other or unspecified Asian | 1.73 | 1.67 - 1.78 | 1.65 | 1.60 - 1.70 |
| Black | 1.15 | 1.12 - 1.19 | 1.14 | 1.10 - 1.17 |
| Mixed | 1.14 | 1.08 - 1.20 | 1.13 | 1.08 - 1.19 |
| Other | 1.48 | 1.41 - 1.56 | 1.44 | 1.37 - 1.51 |
| Unknown | 1.07 | 1.03 - 1.11 | 1.07 | 1.03 - 1.11 |
| **Episodes of sickness absence in 2019** |  |  |  |  |
| 0 | *ref.* | *ref.* | *ref.* | *ref.* |
| 1 | 1.39 | 1.37 - 1.42 | 1.38 | 1.36 - 1.41 |
| 2-3 | 1.83 | 1.79 - 1.86 | 1.80 | 1.77 - 1.84 |
| >3 | 2.41 | 2.36 - 2.47 | 2.38 | 2.32 - 2.43 |
| **Staff group at 9 March 2020** |  |  |  |  |
| Administrative and clerical | *ref.* | *ref.* | *ref.* | *ref.* |
| Additional clinical services | 2.31 | 2.25 - 2.37 | 1.63 | 1.55 - 1.72 |
| Additional professional scientific and technical | 1.37 | 1.31 - 1.43 | 1.05 | 0.98 - 1.12 |
| Allied health professionals | 1.94 | 1.88 - 2.01 | 1.33 | 1.25 - 1.41 |
| Estates and ancillary | 1.45 | 1.39 - 1.50 | 1.30 | 1.25 - 1.35 |
| Healthcare scientists | 1.17 | 1.10 - 1.24 | 1.03 | 0.95 - 1.11 |
| Medical and dental | 1.55 | 1.50 - 1.61 | 1.09 | 1.03 - 1.15 |
| Nursing and midwifery registered | 2.28 | 2.23 - 2.34 | 1.57 | 1.49 - 1.65 |
| Students | 1.87 | 1.60 - 2.20 | 1.35 | 1.14 - 1.59 |
| Multiple or unknown | 1.62 | 1.37 - 1.92 | 1.17 | 0.98 - 1.39 |
| **Exposure category at 9 March 2020\*** |  |  |  |  |
| Care of patients much more likely to have Covid-19 than general population | - | - | 1.48 | 1.40 - 1.57 |
| Care for patients who may be more likely to have Covid-19 than general population | - | - | 1.43 | 1.36 - 1.51 |
| Care of patients with similar or lower prevalence of Covid-19 than general population | - | - | 1.06 | 1.01 - 1.12 |
| No patient care but often in areas where patients have higher prevalence of Covid-19 than general population | - | - | 0.72 | 0.52 - 1.01 |
| No patient care but often in areas where patients have similar or lower prevalence of Covid-19 than general population | - | - | 1.28 | 1.18 - 1.38 |
| No patient care, occasionally in patient areas | - | - | *ref.* | *ref.* |
| Unlikely to be in patient areas, but work with material potentially contaminated by coronavirus | - | - | 0.92 | 0.85 - 0.99 |
| Other or unknown | - | - | 0.73 | 0.71 - 0.76 |

\*exposure categories are based on the constructed JEM. For more information see Supplementary file A.

With no adjustment for exposure category, ORs varied more than twofold across the ten staff groups, the lowest risk being in administrative and clerical jobs (the reference for other risk estimates), and the highest in additional clinical services (OR 2.31), registered nursing and midwifery professionals (OR 2.28), allied health professionals (OR 1.94) and students (OR 1.87). Risk in doctors and dentists was intermediate (OR 1.55), while that in health care scientists was little different from administrative and clerical occupations (OR 1.17).

Exposure category showed an expected gradient of risk, with the highest ORs (relative to no patient care and only occasionally in patient areas) for hands-on or face-to-face care of patients likely to have a higher prevalence of Covid-19 than the general population (Ors 1.48 and 1.43). After adjustment for exposure category, the risk estimates for other staff groups relative to administrative and clerical jobs were all reduced. However, Covid-19 sickness absence was still notably more frequent among those working in additional clinical services (OR 1.63) and in registered nursing and midwifery professionals (OR 1.57). Re-analysis excluding individuals with imputed or missing data gave similar results (Supplementary File C – Table C1).

Most (75%) of the 191 trusts had cared for at least three documented Covid-19 patients by 12 April 2020, but 25 (12.5%) had still not done so by 31.07.2020. The latter were mainly mental health and specialist (e.g. orthopaedic) trusts. Before trusts had cared for three documented Covid-19 patients, ORs for Covid-19 sickness absence relative to administrative and clerical workers were highest in additional clinical services (1.85), registered nurses and midwives (1.81), doctors and dentists (1.66) and allied health professionals (1.62) (Table 3). After trusts had started to care for Covid-19 patients, the ranking of risks by staff group was broadly similar, but the divergence of ORs was greater (2.71 for additional clinical services and 2.70 for registered nurses and midwives). For doctors and dentists, the OR was somewhat reduced (1.45).

**Table 3.**
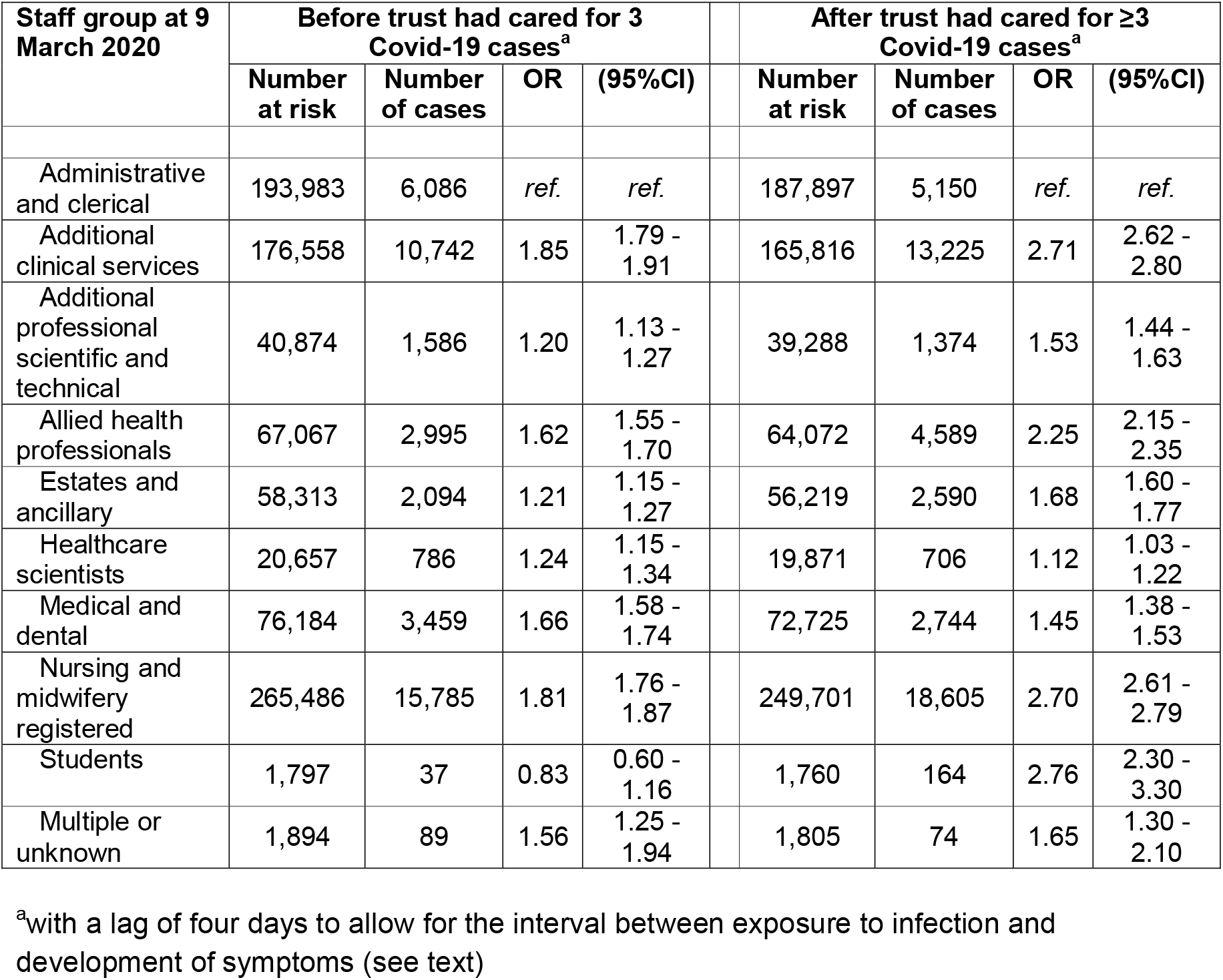
Associations of staff group with a first episode of Covid-19 sickness absence during 9 March to 31 July 2020, according to whether the employing trust had yet cared for at least three documented Covid-19 patients. Risk estimates were derived from two logistic regression models, each of which included all of the variables from Model 1 in Table 2.

Table 4 presents an analysis similar to that in Table 2, but with prolonged Covid-19 sickness absence as the outcome. Individuals with only shorter durations of Covid19 sickness absence were excluded, and risk estimates are relative to no Covid-19 sickness absence. Notable differences from the findings for all Covid-19 sickness absence were a progressive increase in risk across age bands (OR for age >60 vs.<30 years 2.15 in fully adjusted model), higher ORs for non-white vs. white ethnicity, higher risk estimates for additional clinical services and registered nurses and midwives (ORs of 2.88 and 2.59 respectively, reducing to 1.88 and 1.60 after adjustment for exposure category), and lower risk estimates for medical and dental staff (ORs 1.10 and 0.77 before and after adjustment for exposure category).

**Table 4.**
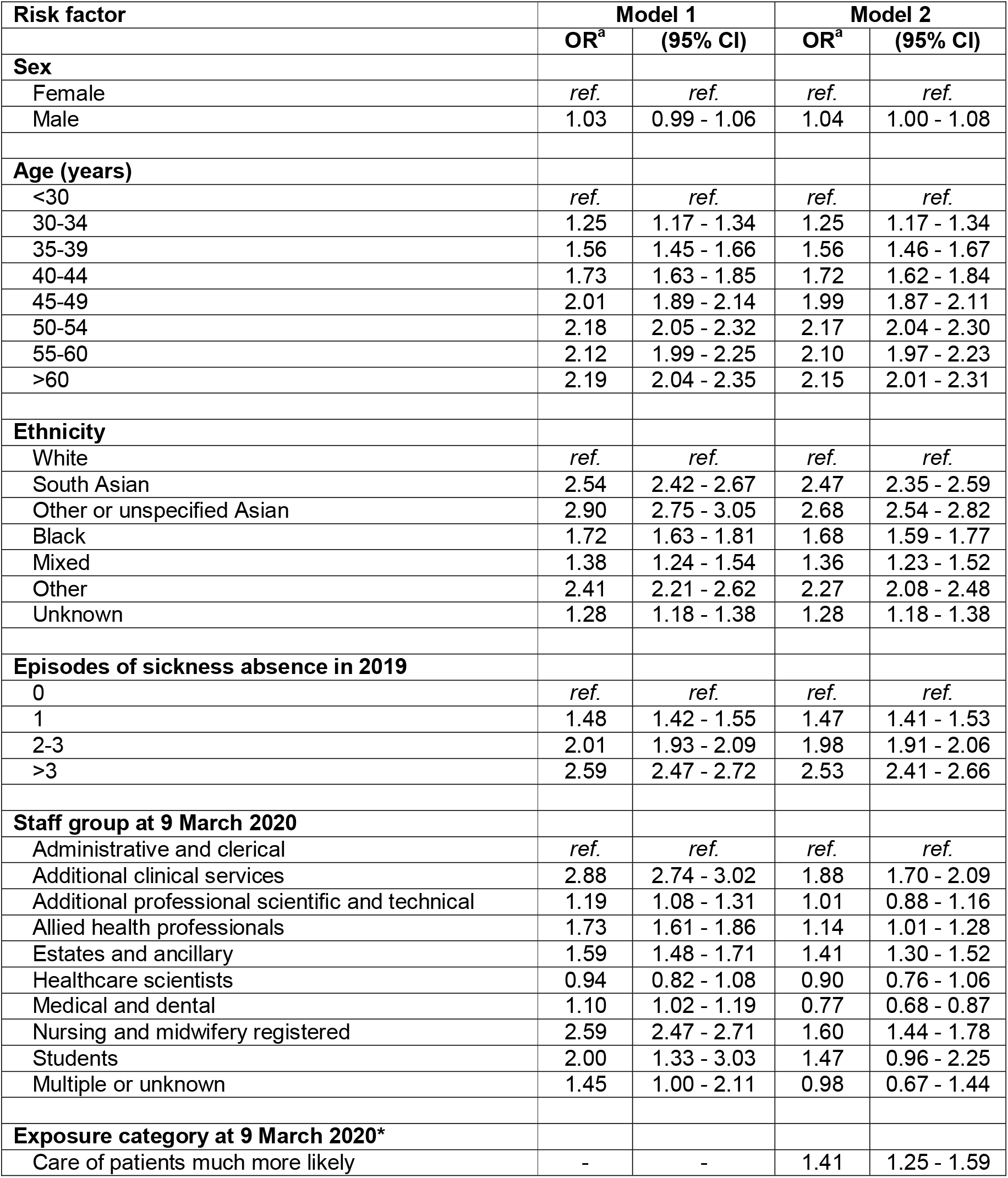
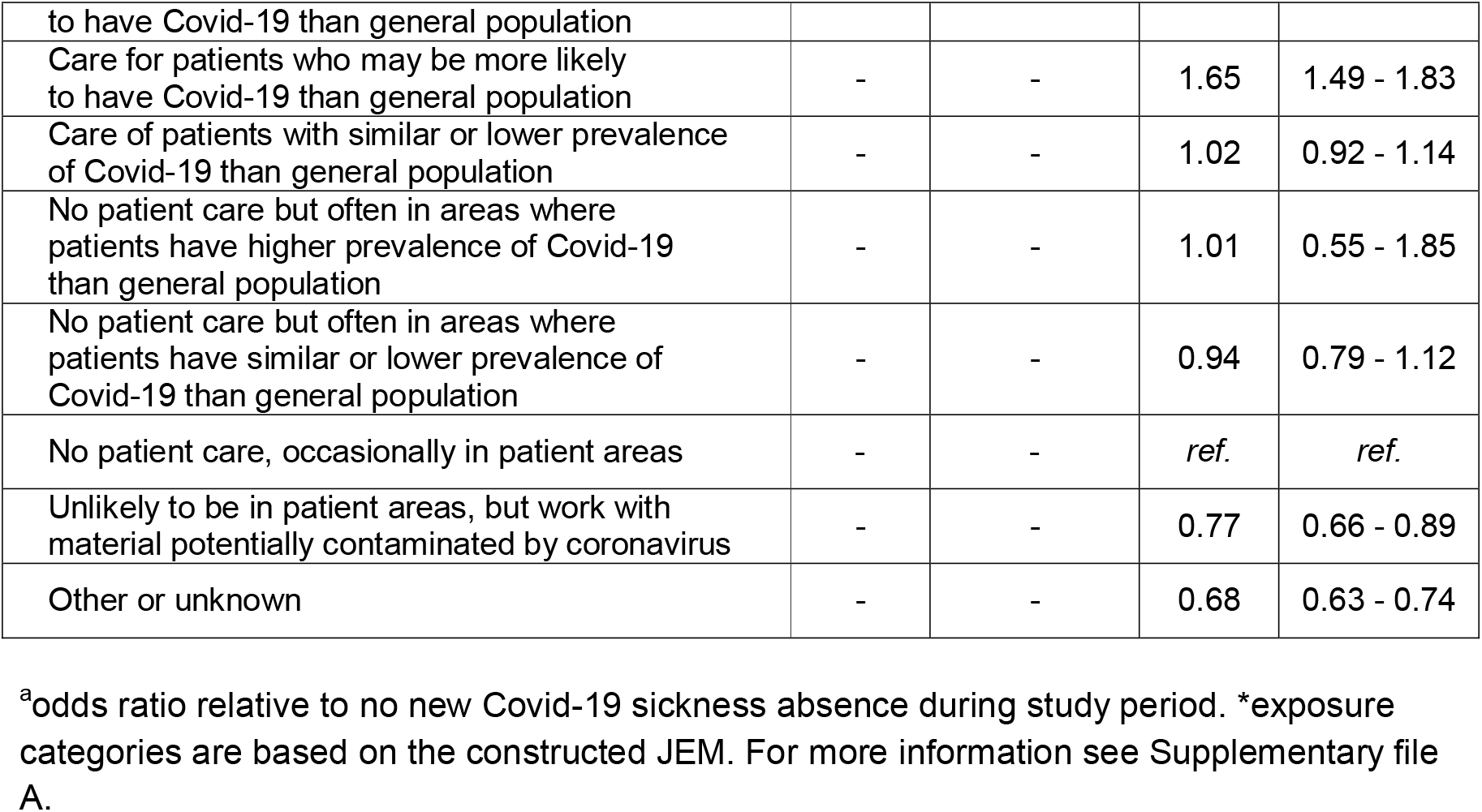
Associations of risk factors at baseline with start of any episode of prolonged Covid-19 sickness absence during 9 March to 16 July 2020. Risk estimates were derived from two logistic regression models that included all of the variables for which results are presented, together with trust (191 categories). An episode of Covid-19 sickness absence was classed as prolonged if it lasted >14 days. Individuals who had only short-term Covid-19 sickness absence were excluded from these analyses (see text).

At the two collaborating trusts, results from antibody tests performed by 7 August 2020 were available for 11,050 staff members. The overall prevalence of positive results among those who had taken Covid-19 sickness absence (37.0%) was 3.3 times that in those who had not (11.1%). There were no differences in this ratio by staff group that could not easily be attributable to random sampling variation (Table 5).

**Table 5.** Results of antibody tests at two trusts according to risk factors. Antibody test results, prior to 7 August 2020, were provided by Cambridge University Hospitals NHS Foundation Trust and Guys and St Thomas’s Trust.

| Risk factor | No Covid-19 sickness absence |  |  |  | Covid-19 sickness absence |  |  |  | Ratio of proportions with at least one positive test<sup>c</sup> |
| --- | --- | --- | --- | --- | --- | --- | --- | --- | --- |
|  | At least one test performed |  | At least one test positive |  | At least one test performed |  | At least one test positive |  |  |
|  | N | (%)<sup>a</sup> | N | (%)<sup>b</sup> | N | (%)<sup>a</sup> | N | (%)<sup>b</sup> |  |
| All employees | 9,502 | 54.8 | 1,053 | 11.1 | 1,548 | 64.6 | 573 | 37.0 | 3.3 |
| Staff group at 9 March 2020 |  |  |  |  |  |  |  |  |  |
| Administrative and clerical | 2,120 | 49.6 | 205 | 9.7 | 198 | 55.8 | 72 | 36.4 | 3.8 |
| Additional clinical services | 981 | 51.6 | 126 | 12.8 | 175 | 57.8 | 66 | 37.7 | 2.9 |
| Additional professional scientific and technical | 479 | 66.3 | 45 | 9.4 | 51 | 68.0 | 13 | 25.5 | 2.7 |
| Allied health professionals | 725 | 63.3 | 73 | 10.1 | 137 | 76.5 | 44 | 32.1 | 3.2 |
| Estates and ancillary | 537 | 46.2 | 135 | 25.1 | 112 | 57.4 | 58 | 51.8 | 2.1 |
| Healthcare scientists | 357 | 55.9 | 18 | 5.0 | 32 | 66.7 | 11 | 34.4 | 6.8 |
| Medical and dental | 1,076 | 54.4 | 78 | 7.2 | 129 | 55.6 | 44 | 34.1 | 4.7 |
| Nursing and midwifery registered | 3,202 | 58.5 | 370 | 11.6 | 712 | 70.7 | 265 | 37.2 | 3.2 |
| Students | 7 | 87.5 | 3 | 42.9 | 0 | 0 | 0 | 0 | 0 |
| Multiple or unknown | 18 | 72.0 | 0 | 0 | 2 | 100 | 0 | 0 | - |

<sup>a</sup>prevalence % of having at least one test.
<sup>b</sup>prevalence % among those tested
<sup>c</sup>proportion among those with Covid-19 sickness absence / proportion among those with no Covid-19 sickness absence

## Discussion

After allowance for employing trust, demographic characteristics, and previous frequency of sickness absence, we found more than twofold variation in the risk of Covid-19 sickness absence across major NHS staff groups in England. Differences were reduced, but not eliminated, following adjustment for potential exposure to infected patients or materials, assessed by a JEM. For prolonged Covid-19 sickness absence, the variation in risk was greater.

The analysis benefitted from a large sample size, giving high statistical power, and from its use of data collected prospectively in a standardised format. Information about employing trust, sex, age, staff group and frequency of earlier sickness absence should all have been highly reliable, and we would not expect serious misclassification between the specified categories of ethnicity. A limitation was that staff group distinguished only broad categories of work. Ideally, analysis would have discriminated between occupations in finer detail, but access to that level of information was precluded by data protection rules. We therefore constructed a JEM to group the 659 occupations in the ESR database to eight exposure categories.

As an indicator of occupational exposure to infection from patients, the JEM should have been superior to staff group. For example, within medical and dental personnel, it distinguished specialists in intensive care, expected to have high exposure to patients with Covid-19, from orthopaedic surgeons, whose patients would be expected to have lower prevalence of the disease. However, even in the detailed occupational classification to which the JEM was applied, some job categories were heterogeneous (e.g. nurses in medical wards could not be distinguished from those working in surgery). Moreover, it did not allow for changes in duties during the epidemic, or for use of PPE and its effectiveness. In early April 2020, workers with a long-term condition such as asthma, were advised by Government that they should ‘shield’ and either work from home or not work at all. The health-related characteristics that prompted advice to shield are associated with higher risk of severe outcomes (vulnerability) should an individual contract Covid-19, but not with a higher risk of contracting infection. To bias associations of staff group with sickness absence for Covid-19 importantly, shielding would need to have been substantially more prevalent in some occupational groups than others. This seems unlikely, but if anything, redeployment out of patient-facing roles would be expected to reduce risk estimates for patient-facing occupations.

Thus, the observed associations with the two highest JEM categories, even after adjustment for staff group, support its validity. However, the varying specificity of occupational categories in the JEM complicates interpretation of numerical estimates of risk for exposure levels. Also, the heterogeneous mix of occupations in individual exposure categories, makes it harder to assess the potential for confounding by non-occupational exposures. For these reasons, we focused principally on risk by staff group (a well-established classification of jobs), and used exposure category to help understand the extent to which associations with staff group reflected patient-related exposures..

The other major limitation was the incomplete validity of sickness absence as a marker for Covid-19. Early in the epidemic, diagnostic tests were not widely available, and clinical diagnoses may not have been accurate. Nevertheless, at the trusts which provided data, antibody tests were more than three times as likely to be positive among individuals who had taken Covid-19 sickness absence.

In assessing relative risks by staff group, we adjusted for demographic variables, for trust and for frequency of sickness absence in 2019. The latter was intended as a marker of individual propensity to take sickness absence when ill and showed an expected association with Covid-19 sickness absence. Adjustment for trust was important because rates of infection were known to have varied geographically (10). Moreover, there may have been systematic differences between trusts in the ascertainment and coding of reasons for absence.

In all analyses, we took administrative and clerical workers as the reference for risks in other staff groups. Making up 21.5% of the study sample, they encompassed a range of occupations, including senior managers as well as middle-grade administrative occupations, clerical workers and receptionists. Most will have been office-based, with little or no direct patient contact, and during the epidemic, some may have worked partially or totally from home. Their work may have entailed social contact with colleagues, but not at a level higher than in many occupations outside healthcare. Furthermore, their socio-economic circumstances will have been neither exceptionally good nor poor. Thus, within the demographic strata that we distinguished, their exposures to SARS-CoV-2 should have been representative of the wider working population in their local area.

An indication of differences in risk between staff groups for reasons other than patient-care comes from analysis restricted to the period before each trust began to care for documented Covid-19 cases (Table 3). During that phase, much of the observed variation in risk might be expected to reflect exposure to infection away from work, or through proximity to infected colleagues. However, the highest ORs (between 1.6 and 1.9) were all in patient-facing occupations, suggesting that there may also have been some unrecognised contact with infected patients.

Once trusts were known to be caring for Covid-19 patients, the ORs for most of these occupations were higher, excess relative risks (estimated as OR-1) increasing by 0.6-0.9 (Table 3). An exception were doctors and dentists, in whom ORs were lower when trusts were known to be caring for Covid-19 patients. This may have been because in the early phase of the epidemic, some doctors contracted infection from undiagnosed patients, but that risk of was reduced once testing became more widely available.

Another clue to the impact of patient-related exposures on differences in risk between staff groups is the effect of adjusting risk estimates for exposure category (Table 2). ORs reduced for all staff groups, as expected given a partial correlation between staff group and exposure category. However, the reductions were greatest for patient-facing occupations. For example, the OR for additional clinical services (a group that included care assistants) fell from 2.31 to 1.63, and that for registered nurses and midwives from 2.28 to 1.57. Such changes point strongly to an important contribution from patient-related exposures, but because of the limitations of the JEM, may not have captured them fully.

When allowance is made for the inaccuracy of sickness absence as a marker for disease, and the possibility of a small occupational risk in the reference group of administrative and clerical workers, the results in Tables 2 and 3 suggest that occupational exposures increased the risk of contracting Covid-19 in additional clinical services, registered nurses and midwives, and allied health professionals by a factor of between 1.5 and 2.5. The average relative risk in doctors and dentists appears to have been somewhat lower, but still elevated. Few studies have explored infection rates of Covid-19 in healthcare staff by occupational group during the first wave of infection in England. Zheng, in a study of 1045 staff at a London hospital tested in March/April 2020, found a higher than expected rate of Covid-19 positivity and correspondingly high Covid-19 sickness absence episodes in medical and dental, nursing, midwifery and additional clinical services staff (2). In a study of 11,500 staff at Oxford University Hospitals, tested between March and early June, porters and cleaners had the highest rates of Covid-19 positivity (3).

In our study, it is notable that risk among laboratory scientists was little higher than in administrative and clerical occupations. This suggests that even early in the epidemic, precautions against transmission of SARS-CoV-2 through the handling of clinical samples were effective.

While our main outcome measure was cumulative prevalence of any Covid-19 sickness absence, we also explored risk factors for longer episodes, expecting that prolonged absence might have higher specificity as a marker for Covid-19. Moreover, it would tend to reflect more disabling disease of the type most likely to be considered for compensation. A complication is that it will have depended not only the risk of contracting infection, but also on personal vulnerability once infection occurred. Thus, while risk of any Covid-19 sickness absence was lowest in the oldest age group, that of prolonged absence increased with age (a major determinant of vulnerability (11)). Similarly, the higher risk of prolonged Covid-19 sickness absence among non-white ethnic groups may have been a consequence of higher vulnerability (11). This will be explored further in a separate report.

For most staff groups, ORs were higher for prolonged than for any Covid-19 sickness absence (Table 4), reinforcing the case for a relative risk in the order of two from occupational exposures. The occupational hazard in medical and dental personnel may have been obscured by relatively low vulnerability to severe disease.

Our analysis suggests that during the first wave of the Covid-19 pandemic in England, occupationally-attributable relative risks for Covid-19 among most patient-facing occupations in healthcare workers were in the order of 1.5 to 2.5. For medical and dental personnel, relative risks were a little lower, but still elevated. Better protective measures for these groups should be a priority in the future. Whether relative risks are sufficient to warrant compensation for Covid-19 as an occupational disease in healthcare workers will depend on the regulatory framework, and the required confidence of occupational attribution.

## Data Availability

All data requests should be submitted to the corresponding author for consideration.

## Ethical approval statement

Approval granted by the NHS Health Research Authority (reference 20/SC/0282). The study was registered at ISRCTN: 36352994

## Contributorship statement

All authors contributed to the planning, conduct, analyses and reporting of this manuscript as outlined below.

Diana van der Platt (statistician): was responsible for the statistical aspects of analysis and interpretation of the quantitative aspects of the study.

Ira Madan (Consultant Occupational Physician and Reader): was co-chief investigator with responsibility for advising on study design, analysis and interpretation of results.

David Coggon (Emeritus Professor of Occupational and Environmental Medicine): was responsible for advising on methodological design, analysis and interpretation of results.

Martie van Tongeren (Professor of Occupational and Environmental Medicine): was responsible for advising on study design, analysis and interpretation of results.

Rhiannon Edge (Lecturer in Population Health): was responsible for advising on study design, analysis and interpretation of results.

Rupert Muiry (Research assistant): was responsible for scoping out and reviewing the emerging literature.

Vaughan Parsons (Research manager): was responsible for overseeing the set-up and delivery of the study, and facilitated data collection.

Paul Cullinan (Professor in Occupational and Environmental Respiratory Disease): was chief investigator with responsibility for advising on study design, analysis and interpretation of results. Had overall responsibility for the management and delivery of the study.

## Data Sharing Statement

With permission, source data is available upon request from the NHS Electronic Staff Record (ESR) Warehouse (NHS England)

## Acknowledgments

We are very grateful to the following, without whom the study would not have been possible: Sam Wright, Workforce Information Advisor, NHS Electronic Staff Record, and Mike Vickerman, Workforce Information and Analysis, DHSC. Dr Gavin Debrera (Public Health England) and Dr Kit Harling gave invaluable help in planning the study.

We are grateful too, to Lee Isidore, Manal Sadik and Victoria Thorpe for their helpful input into the interpretation of our findings.

We would also like to thank Cambridge University Hospitals NHS Foundation Trust (Dr Mark Ferris), Guys and St Thomas’s NHS Foundation Trust (Dr Ali Hashtroudi) and Bolton NHS Foundation Trust (Dr Martin Seed) for providing results from their staff antibody testing programmes.

## References

1. Foster P, Neville S. How poor planning left the UK without enough PPE: Financial Times; 202001/03/2021 [Available from: https://www.ft.com/content/9680c20f-7b71-4f65-9bec-0e9554a8e0a7.

2. Iacobucci G. Covid-19: Lack of testing led to patients being discharged to care homes with virus, say auditors. BMJ. 2020;369:m2375.

3. ONS. Coronavirus (COVID-19) related deaths by occupation, England and Wales: deaths registered between 9 March and 28 December 2020. Office for National Statistics; 2021.

4. Zheng C, Hafezi-Bakhtiari N, Cooper V, Davidson H, Habibi M, Riley P, et al. Characteristics and transmission dynamics of COVID-19 in healthcare workers at a London teaching hospital. The Journal of hospital infection. 2020;106(2):325–9.

5. Eyre DW, Lumley SF, O’Donnell D, Campbell M, Sims E, Lawson E, et al. Differential occupational risks to healthcare workers from SARS-CoV-2 observed during a prospective observational study. eLife. 2020;9(e60675).

6. Jones NK, Rivett L, Sparkes D, Forrest S, Sridhar S, Young J, et al. Effective control of SARS-CoV-2 transmission between healthcare workers during a period of diminished community prevalence of COVID-19. eLife. 2020;9(e59391).

7. Timson A, Clair A. Better housing is crucial for our health and the COVID-19 recovery. The Health Foundation; 2020.

8. Beemsterboer W, Stewart R, Groothoff J, Nijhuis F. A literature review on sick leave determinants (1984-2004). International Journal of Occupational Medicine and Environmental Health. 2009;22(2):169–79.

9. NHS England. COVID-19 NHS Situation Report. UK: NHS England; 2021.

10. Kulu H, Dorey P. Infection rates from Covid-19 in Great Britain by geographical units: A model-based estimation from mortality data. Health & Place. 2021;67(102460).

11. Service I. Poverty, Inequality and COVID-19. 2020.

