## Supplementary File A1 for "Occupational risks of COVID-19 in NHS workers in England"

### Supplementary file A

SOURCE MATERIAL AND PREPARATION OF DATA FOR ANALYSIS

### SOURCE MATERIAL

After ethical approval of our study protocol by the National Health Service (NHS) Health Research Authority (reference 20/SC/0282), the NHS Electronic Staff Record (ESR) Central Team prepared two pseudonymised data files to which we were given access.

### SIP File

The first file (“SIP”) comprised records for all individuals who were continuously employed by NHS trusts in England and Wales from 1 January 2019 (or before) to 31 July 2020. Each record had the following fields:

#### Coded identifier

A unique, encrypted 8 to 9 digit serial number, assigned by the ESR Central Team, which would enable linkage to other data (see below)

#### Sex

Two categories

#### Age band

Nine categories defined by the individual’s age at 15^th^ of September 2020.

#### Ethnicity

Sixty categories

#### Trust

Separate categories for each of 200 trusts – individuals were classified according to the trust by which they were employed at 31 July 2020

#### NHS region

The NHS region in which the employing trust was located (nine categories)

#### Staff group

A single variable with 10 categories specified as in Table A1 (further information on this classification can be found at [https://digital.nhs.uk/binaries/content/assets/website-assets/data-and-information/data-sets/nwd-and-nhs-occupation-codes/nwd_v3.0_data_set_specification_v1.0_final.xlsx](https://eur03.safelinks.protection.outlook.com/?url=https%3A%2F%2Fdigital.nhs.uk%2Fbinaries%2Fcontent%2Fassets%2Fwebsite-assets%2Fdata-and-information%2Fdata-sets%2Fnwd-and-nhs-occupation-codes%2Fnwd_v3.0_data_set_specification_v1.0_final.xlsx&data=04%7C01%7Cdnmc%40mrc.soton.ac.uk%7C16ed21d4b2a9484576c608d8a02dd60b%7C4a5378f929f44d3ebe89669d03ada9d8%7C0%7C0%7C637435473606809409%7CUnknown%7CTWFpbGZsb3d8eyJWIjoiMC4wLjAwMDAiLCJQIjoiV2luMzIiLCJBTiI6Ik1haWwiLCJXVCI6Mn0%3D%7C1000&sdata=XIXz6q22%2F7nf5BPbXgiEuTGe1PrXD6MRBOcWILE67Kw%3D&reserved=0)).

Individuals were classed according to their staff group at 31 July 2020.

### Table A1. Specification of staff groups

| **Staff group** | **Definition** | **Example job roles** |
| --- | --- | --- |
| Administrative and clerical | Non-clinical staff including non-clinical managers, administrative officers, executive board members who do not have significant patient contact as part of their role | Accountant, chief executive, clerical worker, receptionist |
| Additional clinical services | Staff directly supporting those in clinical roles. Support to nursing, allied health professionals, health care scientists and other scientific staff are included. Have significant patient contact as part of their role. | Call operator, emergency care assistant, healthcare assistant, nursery nurse |
| Additional professional scientific and technical | Scientific staff including registered pharmacists, psychologists, social workers, and other roles such as technicians and psychological therapists | Pharmacist, chaplain, social worker, osteopath |
| Allied health professionals | Registered clinical staff providing diagnostic, technical and therapeutic patient care, including dietitians, radiographers and physiotherapists. Includes qualified ambulance staff such as paramedics | Dietitian, physiotherapist, paramedic, drama therapist, specialist practitioner |
| Estates and ancillary | Non-clinical support and maintenance staff, including gardeners, plumbers, cooks and housekeepers who do not have significant patient contact as part of their role | Electrician, housekeeper, telephonist |
| Healthcare scientists | Registered qualified and other staff working in a defined healthcare scientist role, including clinical scientists and biomedical scientists and technicians working in healthcare science. Also includes public health scientific staff | Healthcare scientist, consultant healthcare scientist, healthcare science practitioner |
| Medical and dental | Registered doctors and dentists | Consultant, clinical assistant, dental officer, foundation year 1, specialty doctor |
| Nursing and midwifery registered | Registered nurses and midwives | Staff nurse, midwife, community nurse, modern matron, nurse consultant |
| Students | Directly employed staff undertaking formal education, including student nurses and midwives | Student midwife, student dietitian, student orthoptist |
| No staff group specified |  |  |

#### Exposure category

This was a single variable with eight categories (Table A2). Before compilation of the data files, the ESR Management Team provided us with a list of 659 detailed occupational codes, which are part of the National Workforce Data Set (NWD) owned by NHS Digital and are used across workforce data in the NHS. These occupational codes were used to classify occupations in the ESR database. Because of the need to protect privacy, it was not possible to release individual information at this level to the research team. However, based on their personal knowledge, four members of the study team (an occupational hygienist and three senior occupational physicians with long experience of work in the NHS) constructed a job-exposure matrix (JEM) classifying each occupational category to one of the nine exposure categories. The ESR Management Team then applied the JEM to each individual’s detailed occupational code at 31 July 2020, to generate the exposure category in the SIP file. The specificity with which exposures could be assigned, varied by occupation. For example, the occupational coding scheme distinguished doctors working in intensive care from those working in general surgery, but it did not distinguish between nurses in different adult and general services.

### Table A2. Specification of exposure categories

1. Hands-on or face-to-face care of patients much more likely to have Covid-19 than general population
2. Hands-on or face-to-face care of patients who may be more likely to have Covid-19 than general population
3. Hands-on or face-to-face care of patients whose prevalence of Covid-19 is likely to be similar to, or lower than in the general population
4. No hands-on or face-to-face care of patients, but often working in patient areas where patients are more likely to have Covid-19 than general population
5. No hands-on or face-to-face care of patients, but often working in patient areas where the prevalence of Covid-19 among patients is likely to be similar to, or lower than, in the general population
6. No hands-on or face-to-face care of patients, but occasionally in patient areas
7. Unlikely to be in patient areas, but work with material (blood/urine/clothing/equipment/installations) potentially contaminated by virus
8. Other occupation (i.e. not any of 1-7) or occupation unknown

### ABS File

The second (ABS) file included records of all absences during 1 January 2019 to 31 July 2020, other than for annual leave, among staff included in the SIP file. There was one record for each absence, with the following fields:

#### Coded identifier

Specified as for the SIP file

#### Sex

Specified as for the SIP file

#### Age band

Specified as for the SIP file

#### Ethnicity

Specified as for the SIP file

#### Trust

Coded to 200 categories as in the SIP file, except that individuals were classified according to the trust by which they were employed at the date when their absence started

#### NHS region

The NHS region in which the employing trust (at the date when the absence started) was located (nine categories)

#### Staff group

Classified to 10 categories as in the SIP file, but based on the job held at the date when the absence began

#### Exposure category

A single variable with eight categories, specified as in the SIP file, but based on the job held at the date when the absence began

#### Reason for absence

Described by four variables:

- Attendance type (19 categories)
- Absence category (11 categories
- Attendance reason (77 categories)
- Related reason (5 categories)

The categories of the first three of these variables can be found in Table A4 below. The categories of related reason were:

- Coronavirus (COVID-19)
- Coronavirus (COVID-19) – Household Member Symptoms
- Coronavirus (COVID-19) – Post travel Quarantine
- Coronavirus (COVID-19) – Test and Trace Contact
- Menopause

#### Date absence started

Date month and year

#### Date absence ended

Date month and year

#### Numbers of records

In total, 981,131 individuals had at least one record in the SIP file, and of those, 835,180 had at least one recorded absence in the ABS file (no individuals had a record in the ABS file but not in the SIP file). The ABS file contained records on a total of 4,159,991 absence episodes.

### INITIAL STEPS

### Removal of complete duplicates

We first removed all records that were exact duplicates from the SIP file (n = 2,880) and ABS file (53,288).

### Reclassification of ethnicity

The scheme that was used to classify ethnicity in the two data files distinguished 77 different categories. Some were quite rare, limiting the potential for their analysis as separate entities. Also, there were frequent instances of overlap in the specification of categories (e.g. “Indian” with “Asian or Asian British – Indian” and “White English” with White British”).

We therefore aggregated the starting categories into two broader classifications of ethnicity (“Ethnicity group 1” and “Ethnicity group 2”), one nested within the other, with the aim of ensuring adequate numbers for meaningful analysis in each aggregated category, and reducing overlap and ambiguity. Table A3 shows the frequency of each category of ethnicity in the original data files, and the aggregated categories to which it was assigned in the two broader classifications.

### Reclassification of reason for absence

The first three variables that characterised reason for absence (attendance type, absence category, and attendance reason) occurred in 192 different combinations. To facilitate further analysis, these combinations were collapsed into 60 categories of a new variable which we labelled as “Collapsed absence category” (Table A4).

### Table A3. Reclassification of ethnicity

| **Original category** | **Frequency across all SIP and ABS records** | **Allocation in revised classifications** | |
| --- | --- | --- | --- |
|  |  | **Ethnicity group 1** | **Ethnicity group 2** |
| 0 White | 243 | White | White |
| 1 Black-Caribbean | 173 | Black or Black British - Caribbean | Black or Black British |
| 2 Black-African | 911 | Black or Black British - African | Black or Black British |
| 3 Black-Other | 27 | Black or Black British - Any other or unspecified Black background | Black or Black British |
| 4 Indian | 181 | Asian or Asian British - Indian | Asian or Asian British - South Asian |
| 5 Pakistani | 12 | Asian or Asian British - Pakistani | Asian or Asian British - South Asian |
| 7 Chinese | 9 | Asian or Asian British - Any other or unspecified Asian background | Asian or Asian British - Other or unspecified |
| 9 Not given | 47 | Not stated | Not stated |
| A White - British | 3477821 | White | White |
| B White - Irish | 56427 | White | White |
| C White - Any other White background | 188395 | White | White |
| C2 White Northern Irish | 1167 | White | White |
| C3 White Unspecified | 7271 | White | White |
| CA White English | 36682 | White | White |
| CB White Scottish | 3378 | White | White |
| CC White Welsh | 2343 | White | White |
| CD White Cornish | 1686 | White | White |
| CE White Cypriot (non specific) | 206 | White | White |
| CF White Greek | 2157 | White | White |
| CG White Greek Cypriot | 490 | White | White |
| CH White Turkish | 612 | White | White |
| CJ White Turkish Cypriot | 271 | White | White |
| CK White Italian | 5880 | White | White |
| CL White Irish Traveller | 38 | White | White |
| CM White Traveller | 73 | White | White |
| CN White Gypsy/Romany | 251 | White | White |
| CP White Polish | 12075 | White | White |
| CQ White ex-USSR | 873 | White | White |
| CR White Kosovan | 163 | White | White |
| CS White Albanian | 452 | White | White |
| CT White Bosnian | 40 | White | White |
| CU White Croatian | 378 | White | White |
| CV White Serbian | 145 | White | White |
| CW White Other Ex-Yugoslav | 280 | White | White |
| CX White Mixed | 2785 | Mixed | Mixed |
| CY White Other European | 26591 | White | White |
| D Mixed - White & Black Caribbean | 27712 | Mixed | Mixed |
| E Mixed - White & Black African | 13526 | Mixed | Mixed |
| F Mixed - White & Asian | 19742 | Mixed | Mixed |
| G Mixed - Any other mixed background | 23086 | Mixed | Mixed |
| GA Mixed - Black & Asian | 472 | Mixed | Mixed |
| GB Mixed - Black & Chinese | 44 | Mixed | Mixed |
| GC Mixed - Black & White | 1179 | Mixed | Mixed |
| GD Mixed - Chinese & White | 461 | Mixed | Mixed |
| GE Mixed - Asian & Chinese | 440 | Mixed | Mixed |
| GF Mixed - Other/Unspecified | 1706 | Mixed | Mixed |
| H Asian or Asian British - Indian | 238011 | Asian or Asian British - Indian | Asian or Asian British - South Asian |
| J Asian or Asian British - Pakistani | 76282 | Asian or Asian British - Pakistani | Asian or Asian British - South Asian |
| K Asian or Asian British - Bangladeshi | 27145 | Asian or Asian British - Bangladeshi | Asian or Asian British - South Asian |
| L Asian or Asian British - Any other Asian background | 132728 | Asian or Asian British - Any other or unspecified Asian background | Asian or Asian British - Other or unspecified |
| LA Asian Mixed | 1601 | Mixed | Mixed |
| LB Asian Punjabi | 1328 | Asian or Asian British - Indian | Asian or Asian British - South Asian |
| LC Asian Kashmiri | 250 | Asian or Asian British - Indian | Asian or Asian British - South Asian |
| LD Asian East African | 482 | Other | Other |
| LE Asian Sri Lankan | 2468 | Asian or Asian British - Sri Lankan | Asian or Asian British - South Asian |
| LF Asian Tamil | 985 | Asian or Asian British - Indian | Asian or Asian British - South Asian |
| LG Asian Sinhalese | 180 | Asian or Asian British - Sri Lankan | Asian or Asian British - South Asian |
| LH Asian British | 6769 | Asian or Asian British - Any other or unspecified Asian background | Asian or Asian British - Other or unspecified |
| LJ Asian Caribbean | 743 | Other | Other |
| LK Asian Unspecified | 6325 | Asian or Asian British - Any other or unspecified Asian background | Asian or Asian British - Other or unspecified |
| M Black or Black British - Caribbean | 87878 | Black or Black British - Caribbean | Black or Black British |
| N Black or Black British - African | 206147 | Black or Black British - African | Black or Black British |
| Nan | 21209 | Not stated | Not stated |
| P Black or Black British - Any other Black background | 18128 | Black or Black British - Any other or unspecified Black background | Black or Black British |
| PA Black Somali | 2367 | Black or Black British - African | Black or Black British |
| PB Black Mixed | 503 | Mixed | Mixed |
| PC Black Nigerian | 9720 | Black or Black British - African | Black or Black British |
| PD Black British | 11356 | Black or Black British - Any other or unspecified Black background | Black or Black British |
| PE Black Unspecified | 1435 | Black or Black British - Any other or unspecified Black background | Black or Black British |
| R Chinese | 17949 | Asian or Asian British - Any other or unspecified Asian background | Asian or Asian British - Other or unspecified |
| S Any Other Ethnic Group | 64865 | Other | Other |
| SA Vietnamese | 213 | Asian or Asian British - Any other or unspecified Asian background | Asian or Asian British - Other or unspecified |
| SB Japanese | 273 | Asian or Asian British - Any other or unspecified Asian background | Asian or Asian British - Other or unspecified |
| SC Filipino | 55696 | Asian or Asian British - Any other or unspecified Asian background | Asian or Asian British - Other or unspecified |
| SD Malaysian | 705 | Asian or Asian British - Any other or unspecified Asian background | Asian or Asian British - Other or unspecified |
| SE Other Specified | 6487 | Other | Other |
| Z Not Stated | 169316 | Not stated | Not stated |
| Missing | 4056 | Not stated | Not stated |

**Table A4.	Specification of collapsed absence categories**

| **Attendance Type** | **Absence Category** | **Attendance Reason** | **Collapsed absence category** |
| --- | --- | --- | --- |
| Paid Part Day | Paid Leave | Adoption Appointment | Adoption appointment |
| Special Decreasing Bal | Special Leave | Adoption Appointment | Adoption appointment |
| Special Increasing Bal | Special Leave | Adoption Appointment | Adoption appointment |
| Unpaid Authorised Special | Special Leave | Adoption Appointment | Adoption appointment |
| Unpaid Authorised Special Hrs | Special Leave | Adoption Appointment | Adoption appointment |
| Adoption | Adoption | Adoption Leave | Adoption leave |
| Adoption | Adoption |  | Adoption leave |
| Unpaid Authorised Special | Special Leave | Adoption Leave | Adoption leave |
| Unpaid Authorised Special Hrs | Special Leave | Adoption Leave | Adoption leave |
| Paternity Adoption | Paternity Adoption | Paternity Leave | Adoption leave paternity |
| Paternity Adoption | Paternity Adoption |  | Adoption leave paternity |
| Shared Parental Adoption | Shared Parental Adoption |  | Adoption leave shared parental adoption |
| Unpaid Authorised Special Hrs | Special Leave | Annual Leave | Annual leave |
| Special Decreasing Bal | Special Leave | Antenatal | Antenatal |
| Special Increasing Bal | Special Leave | Antenatal | Antenatal |
| Paid Part Day | Paid Leave | Attendance at Public Bodies | Attendance public body |
| Special Decreasing Bal | Special Leave | Attendance at Public Bodies | Attendance public body |
| Special Increasing Bal | Special Leave | Attendance at Public Bodies | Attendance public body |
| Unpaid Authorised Special | Special Leave | Attendance at Public Bodies | Attendance public body |
| Unpaid Authorised Special Hrs | Special Leave | Attendance at Public Bodies | Attendance public body |
| Special Decreasing Bal | Special Leave | Bereavement | Bereavement |
| Special Increasing Bal | Special Leave | Bereavement | Bereavement |
| Unpaid Authorised Special | Special Leave | Bereavement | Bereavement |
| Unpaid Authorised Special Hrs | Special Leave | Bereavement | Bereavement |
| Special Decreasing Bal | Special Leave | Career Break | Career break |
| Special Increasing Bal | Special Leave | Career Break | Career break |
| Unpaid Authorised Special | Special Leave | Career Break | Career break |
| Unpaid Authorised Special Hrs | Special Leave | Career Break | Career break |
| Paid Part Day | Paid Leave | Carer's Leave | Carer's leave |
| Special Decreasing Bal | Special Leave | Carer's Leave | Carer's leave |
| Special Decreasing Bal | Special Leave | Emergency Leave/Time Off for Dependants | Carer's leave |
| Special Increasing Bal | Special Leave | Carer's Leave | Carer's leave |
| Special Increasing Bal | Special Leave | Emergency Leave/Time Off for Dependants | Carer's leave |
| Unpaid Authorised Special | Special Leave | Carer's Leave | Carer's leave |
| Unpaid Authorised Special | Special Leave | Emergency Leave/Time Off for Dependants | Carer's leave |
| Unpaid Authorised Special Hrs | Special Leave | Carer's Leave | Carer's leave |
| Unpaid Authorised Special Hrs | Special Leave | Emergency Leave/Time Off for Dependants | Carer's leave |
| Paid Part Day | Paid Leave | Compassionate Leave | Compassionate leave |
| Special Decreasing Bal | Special Leave | Compassionate Leave | Compassionate leave |
| Special Increasing Bal | Special Leave | Compassionate Leave | Compassionate leave |
| Unpaid Authorised Special | Special Leave | Compassionate Leave | Compassionate leave |
| Unpaid Authorised Special Hrs | Special Leave | Compassionate Leave | Compassionate leave |
| Special Decreasing Bal | Special Leave | Court Appearance | Court appearance |
| Special Increasing Bal | Special Leave | Court Appearance | Court appearance |
| Unpaid Authorised Special | Special Leave | Court Appearance | Court appearance |
| Unpaid Authorised Special Hrs | Special Leave | Court Appearance | Court appearance |
| Paid Part Day | Paid Leave | Disability Leave | Disability leave |
| Special Decreasing Bal | Special Leave | Disability Leave | Disability leave |
| Special Increasing Bal | Special Leave | Disability Leave | Disability leave |
| Unpaid Authorised Special | Special Leave | Disability Leave | Disability leave |
| Unpaid Authorised Special Hrs | Special Leave | Disability Leave | Disability leave |
| Paid Part Day | Paid Leave | Industrial Action | Industrial action |
| Special Increasing Bal | Special Leave | Industrial Action | Industrial action |
| Unpaid Authorised Special | Special Leave | Industrial Action | Industrial action |
| Unpaid Authorised Special Hrs | Special Leave | Industrial Action | Industrial action |
| Unpaid Unauth Special Hrs | Special Leave | Industrial Action | Industrial action |
| Unpaid Unauthorised Special | Special Leave | Industrial Action | Industrial action |
| Special Decreasing Bal | Special Leave | Infection Precaution | Infection precaution |
| Special Increasing Bal | Special Leave | Infection Precaution | Infection precaution |
| Paid Part Day | Paid Leave | Interview Leave | Interview leave |
| Special Decreasing Bal | Special Leave | Interview Leave | Interview leave |
| Special Increasing Bal | Special Leave | Interview Leave | Interview leave |
| Unpaid Authorised Special | Special Leave | Interview Leave | Interview leave |
| Unpaid Authorised Special Hrs | Special Leave | Interview Leave | Interview leave |
| Special Decreasing Bal | Special Leave | Jury Service | Jury service |
| Special Increasing Bal | Special Leave | Jury Service | Jury service |
| Unpaid Authorised Special | Special Leave | Jury Service | Jury service |
| Unpaid Authorised Special Hrs | Special Leave | Jury Service | Jury service |
| Maternity | Maternity | Maternity Leave | Maternity leave |
| Maternity | Maternity |  | Maternity leave |
| Medical Suspension with Pay | Paid Leave | Allergy | Medical suspension |
| Medical Suspension with Pay | Paid Leave | Infection | Medical suspension |
| Medical Suspension with Pay | Paid Leave | Needlestick | Medical suspension |
| Medical Suspension with Pay | Paid Leave | Other | Medical suspension |
| Medical Suspension with Pay | Paid Leave |  | Medical suspension |
| Special Decreasing Bal | Special Leave | Medical Suspension | Medical suspension |
| Special Increasing Bal | Special Leave | Medical Suspension | Medical suspension |
| Paid Part Day | Paid Leave | Medical/Dental Appointment | Medical/dental appointment |
| Special Decreasing Bal | Special Leave | Medical/Dental Appointment | Medical/dental appointment |
| Special Increasing Bal | Special Leave | Medical/Dental Appointment | Medical/dental appointment |
| Unpaid Authorised Special | Special Leave | Medical/Dental Appointment | Medical/dental appointment |
| Unpaid Authorised Special Hrs | Special Leave | Medical/Dental Appointment | Medical/dental appointment |
| Paid Part Day | Paid Leave | Other | Other part day |
| Paid Part Day | Paid Leave | Paid Part Day | Other part day |
| Paid Part Day | Paid Leave | Time Off in Lieu - Other | Other part day |
| Paid Part Day | Paid Leave | Time Off in Lieu - Overtime/Time Owed | Other part day |
| Paid Part Day | Paid Leave | Time Off in Lieu - Worked Public Holiday | Other part day |
| Paid Part Day | Paid Leave | Trade Union Duties | Other part day |
| Paid Part Day | Paid Leave |  | Other part day |
| Special Decreasing Bal | Special Leave | Other | Other special leave |
| Special Decreasing Bal | Special Leave | Time Off in Lieu - Other | Other special leave |
| Special Decreasing Bal | Special Leave | Time Off in Lieu - Overtime/Time Owed | Other special leave |
| Special Decreasing Bal | Special Leave | Time Off in Lieu - Worked Public Holiday | Other special leave |
| Special Decreasing Bal | Special Leave | Trade Union Duties | Other special leave |
| Special Decreasing Bal | Special Leave |  | Other special leave |
| Special Increasing Bal | Special Leave | Other | Other special leave |
| Special Increasing Bal | Special Leave |  | Other special leave |
| Unpaid Authorised Special | Special Leave | Other | Other special leave |
| Unpaid Authorised Special | Special Leave |  | Other special leave |
| Unpaid Authorised Special Hrs | Special Leave | Other | Other special leave |
| Unpaid Authorised Special Hrs | Special Leave |  | Other special leave |
| Unpaid Unauth Special Hrs | Special Leave | Other | Other special leave |
| Unpaid Unauth Special Hrs | Special Leave | Unauthorised Leave | Other special leave |
| Unpaid Unauth Special Hrs | Special Leave |  | Other special leave |
| Unpaid Unauthorised Special | Special Leave | Other | Other special leave |
| Unpaid Unauthorised Special | Special Leave | Unauthorised Leave | Other special leave |
| Unpaid Unauthorised Special | Special Leave |  | Other special leave |
| Special Increasing Bal | Special Leave | Time Off in Lieu - Other | Other special leave time off in lieu |
| Special Increasing Bal | Special Leave | Time Off in Lieu - Overtime/Time Owed | Other special leave time off in lieu |
| Special Increasing Bal | Special Leave | Time Off in Lieu - Worked Public Holiday | Other special leave time off in lieu |
| Special Increasing Bal | Special Leave | Trade Union Duties | Other special leave trade union |
| Unpaid Authorised Special | Special Leave | Trade Union Duties | Other special leave trade union |
| Special Decreasing Bal | Special Leave | Parental Leave | Parental leave |
| Special Increasing Bal | Special Leave | Parental Leave | Parental leave |
| Unpaid Authorised Special | Special Leave | Parental Leave | Parental leave |
| Unpaid Authorised Special Hrs | Special Leave | Parental Leave | Parental leave |
| Shared Parental Birth | Shared Parental Birth |  | Parental leave shared parental birth |
| Paid Part Day | Paid Leave | Paternity Antenatal Appointment | Paternity appointment |
| Special Decreasing Bal | Special Leave | Paternity Antenatal Appointment | Paternity appointment |
| Special Increasing Bal | Special Leave | Paternity Antenatal Appointment | Paternity appointment |
| Unpaid Authorised Special | Special Leave | Paternity Antenatal Appointment | Paternity appointment |
| Unpaid Authorised Special Hrs | Special Leave | Paternity Antenatal Appointment | Paternity appointment |
| Additional Pat Leave Birth | Additional Paternity Birth |  | Paternity leave |
| Paternity Birth | Paternity Birth | Paternity Leave | Paternity leave |
| Paternity Birth | Paternity Birth |  | Paternity leave |
| Unpaid Authorised Special | Special Leave | Paternity Leave | Paternity leave |
| Unpaid Authorised Special Hrs | Special Leave | Paternity Leave | Paternity leave |
| Paid Part Day | Paid Leave | Phased Return to Work | Phased return |
| Special Decreasing Bal | Special Leave | Phased Return to Work | Phased return |
| Special Increasing Bal | Special Leave | Phased Return to Work | Phased return |
| Unpaid Authorised Special | Special Leave | Phased Return to Work | Phased return |
| Unpaid Authorised Special Hrs | Special Leave | Phased Return to Work | Phased return |
| Special Decreasing Bal | Special Leave | Magisterial/Local Government/Parliamentary Candidate | Political candidate |
| Special Increasing Bal | Special Leave | Magisterial/Local Government/Parliamentary Candidate | Political candidate |
| Unpaid Authorised Special | Special Leave | Magisterial/Local Government/Parliamentary Candidate | Political candidate |
| Unpaid Authorised Special Hrs | Special Leave | Magisterial/Local Government/Parliamentary Candidate | Political candidate |
| Sickness | Sickness | S14 Asthma | Sickness asthma |
| Sickness | Sickness | S11 Back Problems | Sickness back problems |
| Sickness | Sickness | S18 Blood disorders | Sickness blood disorders |
| Sickness | Sickness | S20 Burns, poisoning, frostbite, hypothermia | Sickness burns, poisoning, frostbite, hypothermia |
| Sickness | Sickness | S17 Benign and malignant tumours, cancers | Sickness cancer |
| Sickness | Sickness | S19 Heart, cardiac & circulatory problems | Sickness cardiac and circulatory |
| Sickness | Sickness | S15 Chest & respiratory problems | Sickness chest and respiratory |
| Sickness | Sickness | S13 Cold, Cough, Flu - Influenza | Sickness cough, flu |
| Sickness | Sickness | S22 Dental and oral problems | Sickness dental and oral problems |
| Sickness | Sickness | S21 Ear, nose, throat (ENT) | Sickness ear, nose, throat |
| Sickness | Sickness | S24 Endocrine / glandular problems | Sickness endocrine, glandular problems |
| Sickness | Sickness | S23 Eye problems | Sickness eye problems |
| Sickness | Sickness | S25 Gastrointestinal problems | Sickness gastrointestinal problems |
| Sickness | Sickness | S26 Genitourinary & gynaecological disorders | Sickness genitourinary, gynaecological problems |
| Sickness | Sickness | S16 Headache / migraine | Sickness headache, migraine |
| Sickness | Sickness | S27 Infectious diseases | Sickness infectious diseases |
| Sickness | Sickness | S28 Injury, fracture | Sickness injury, fracture |
| Sickness | Sickness | S10 Anxiety/stress/depression/other psychiatric illnesses | Sickness mental health |
| Sickness | Sickness | Stress | Sickness mental health |
| Sickness | Sickness | S29 Nervous system disorders | Sickness nervous system disorders |
| Sickness | Sickness | S98 Other known causes - not elsewhere classified | Sickness other |
| Sickness | Sickness | Musculo-skeletal Back | Sickness other MSDs |
| Sickness | Sickness | Musculo-skeletal Other Joint, Lower Limb | Sickness other MSDs |
| Sickness | Sickness | S12 Other musculoskeletal problems | Sickness other MSDs |
| Paid Part Day | Paid Leave | Sickness | Sickness part day |
| Sickness | Sickness | Pregnancy Related | Sickness pregnancy related disorders |
| Sickness | Sickness | S30 Pregnancy related disorders | Sickness pregnancy related disorders |
| Sickness | Sickness | S31 Skin disorders | Sickness skin disorders |
| Sickness | Sickness | S32 Substance abuse | Sickness substance abuse |
| Sickness | Sickness | Surgery | Sickness surgery |
| Sickness | Sickness | Not Known | Sickness unknow cause |
| Sickness | Sickness | S99 Unknown causes / Not specified | Sickness unknow cause |
| Study Decreasing Bal | Study Leave | Professional Leave | Study leave |
| Study Decreasing Bal | Study Leave | Study Leave | Study leave |
| Study Decreasing Bal | Study Leave |  | Study leave |
| Study Increasing Bal | Study Leave | Study Leave | Study leave |
| Study Increasing Bal | Study Leave |  | Study leave |
| Training Development | Paid Leave | Development | Study leave |
| Training Development | Paid Leave | External Training | Study leave |
| Training Development | Paid Leave | Internal Training | Study leave |
| Training Development | Paid Leave | Other | Study leave |
| Training Development | Paid Leave |  | Study leave |
| Special Decreasing Bal | Special Leave | Suspended - Paid | Suspended |
| Special Increasing Bal | Special Leave | Suspended - Paid | Suspended |
| Unpaid Authorised Special | Special Leave | Suspended - Unpaid | Suspended |
| Unpaid Authorised Special Hrs | Special Leave | Suspended - Unpaid | Suspended |
| Special Decreasing Bal | Special Leave | Deployment with Reserve Forces | Volunteer |
| Special Decreasing Bal | Special Leave | Training with Reserve and Cadet Forces | Volunteer |
| Special Decreasing Bal | Special Leave | Volunteer Leave | Volunteer |
| Special Increasing Bal | Special Leave | Deployment with Reserve Forces | Volunteer |
| Special Increasing Bal | Special Leave | Training with Reserve and Cadet Forces | Volunteer |
| Special Increasing Bal | Special Leave | Volunteer Leave | Volunteer |
| Unpaid Authorised Special | Special Leave | Training with Reserve and Cadet Forces | Volunteer |
| Unpaid Authorised Special | Special Leave | Volunteer Leave | Volunteer |
| Unpaid Authorised Special Hrs | Special Leave | Training with Reserve and Cadet Forces | Volunteer |
| Unpaid Authorised Special Hrs | Special Leave | Volunteer Leave | Volunteer |

### INCONSISTENCIES AND ANOMALIES IN DATA ON INDIVIDUAL ABSENCE EPISODES

We next considered each individual episode of absence, looking for inconsistencies and anomalies in the recorded data. These took several forms.

### Inconsistent dates of absence

Two episodes, both for maternity leave, had an end date before their start date. In both cases, we assumed that the record was corrupted as it is not possible for a user to enter an end date that is prior to the start date, and therefore we changed the incorrect end date to one year later. This meant that the duration of the leave in each case was a little less than 12 months, which was a common duration for maternity leave in the dataset as a whole. In one of the two cases, there was also an absence for “medical suspension” linked to Covid-19 which started four weeks after the maternity leave. However, such overlap occurred quite frequently with maternity leave and was not judged to invalidate the imputed end date.

### Implausible durations of absence

18 end dates, all of which were after 31 July 2020, gave implausibly long durations that were incompatible with the reason for absence. These appeared to have arisen from transposition of digits in data entry (e.g. 2019 being entered as 2109 or 2018 being entered as 2028), and were revised accordingly.

### Missing end dates for absences

The approach to missing end dates depended on whether there was another episode of absence with a later start date.

Where there was no subsequent episode with a later start date, we assumed that an absence continued beyond 31 July 2020, provided that would not make it implausibly long (n = 60,805). In this context, plausibility was determined from the distribution of durations for all absences in the same collapsed absence category with known start and end dates. Where continuation beyond 31 July 2020 would imply an implausibly long duration, we imputed an end date by adding the median duration for the collapsed absence category to the start date (n = 466).

Where the individual had another episode with a later start date, the missing end date was imputed as the earlier of: a) the start date of the episode plus the median duration for the collapsed absence category; and b) the start date of the next episode for that individual minus one day (n = 6,938).

### ELIMINATION OF ABSENCES THAT ENDED BEFORE THE START OF THE STUDY PERIOD

Having made these adjustments, we removed from the ABS file, all records for absences that ended before 01 January 2019 (n = 1,567), leaving a total of 4,103,602 remaining absence records.

### OTHER INCONSISTENCIES AND ANOMALIES

Where an individual had one or more records in the ABS file as well as a record in the SIP file, it was important to check that all of his/her records were mutually consistent. Some characteristics (sex, age group and ethnicity) could not change over time and should therefore have been identical in all records for the same individual. Others (trust, staff group, exposure category) were specific to the date of the record (31 July 2020 for the SIP file and date absence started for the ABS file) and might have changed over time if people changed jobs. Furthermore, it was possible that some individuals held more than one job simultaneously, and that trust, staff group and/or exposure category differed between those jobs. It was also important to check that for each individual, the start and end dates of all absence episodes were mutually compatible.

When exploring inconsistencies, we considered the possibility that records might have been assigned to the wrong individual. A pointer to this would be inconsistencies for multiple demographic variables in the same person. However, only one individual had inconsistencies in both of sex and ethnicity, one in both age group and ethnicity, and none in both sex and age group. We therefore judged such error to be unlikely.

### Sex

Sex was recorded inconsistently in 33 individuals. This may have occurred because first names were not clearly sex-specific. Each individual was assigned the modal value for sex across all of his/her records, giving preference to the value from the SIP file in the event of a tie.

### Age group

Age group was recorded inconsistently in 45 individuals. Each was assigned the modal value for age group across all of his/her records, giving preference to the value from the SIP file in the event of a tie.

### Ethnicity

Across all of their records in the SIP and ABS files, 6,872 individuals were assigned to two or three different categories of ethnicity. In many cases this reflected missing information or a lack of specificity in some records as compared with others. Table A5 shows the conventions that we adopted in reclassifying each combination of ethnicity categories to a single category in Ethnicity group 1 and Ethnicity group 2. Table A5 also shows the frequency with which each combination occurred in the dataset.

**Table A5.	Assignment of Ethnicity groups 1 and 2 where more than one category of ethnicity was assigned to the same individual in the SIP and ABS data files**

| **Combination of different ethnicity categories for same individual** | | | **Number of individuals** | **Allocation in revised classifications** | |
| --- | --- | --- | --- | --- | --- |
|  |  |  |  | **Ethnicity group 1** | **Ethnicity group 2** |
| White | Asian or Asian British - Any other or unspecified Asian background |  | 28 | Mixed | Mixed |
| White | Asian or Asian British - Bangladeshi |  | 6 | Mixed | Mixed |
| White | Asian or Asian British - Indian |  | 37 | Mixed | Mixed |
| White | Asian or Asian British - Pakistani |  | 15 | Mixed | Mixed |
| White | Black or Black British - African |  | 11 | Mixed | Mixed |
| White | Black or Black British - Any other or unspecified Black background |  | 6 | Mixed | Mixed |
| White | Black or Black British - Caribbean |  | 26 | Mixed | Mixed |
| White | Mixed | Not stated | 1 | Mixed | Mixed |
| White | Mixed |  | 291 | Mixed | Mixed |
| White | Not stated |  | 3675 | White | White |
| White | Other | Not stated | 1 | Mixed | Mixed |
| White | Other |  | 124 | Mixed | Mixed |
| Asian or Asian British - Any other or unspecified Asian background | Asian or Asian British - Bangladeshi |  | 16 | Asian or Asian British - Bangladeshi | Asian or Asian British - South Asian |
| Asian or Asian British - Any other or unspecified Asian background | Asian or Asian British - Indian |  | 103 | Asian or Asian British - Indian | Asian or Asian British - South Asian |
| Asian or Asian British - Any other or unspecified Asian background | Asian or Asian British - Pakistani |  | 30 | Asian or Asian British - Pakistani | Asian or Asian British - South Asian |
| Asian or Asian British - Any other or unspecified Asian background | Asian or Asian British - Sri Lankan |  | 11 | Asian or Asian British - Sri Lankan | Asian or Asian British - South Asian |
| Asian or Asian British - Any other or unspecified Asian background | Black or Black British - African |  | 3 | Mixed | Mixed |
| Asian or Asian British - Any other or unspecified Asian background | Black or Black British - Any other or unspecified Black background |  | 6 | Mixed | Mixed |
| Asian or Asian British - Any other or unspecified Asian background | Not stated |  | 302 | Asian or Asian British - Any other or unspecified Asian background | Asian or Asian British - Other or unspecified |
| Asian or Asian British - Any other or unspecified Asian background | Other |  | 374 | Asian or Asian British - Any other or unspecified Asian background | Asian or Asian British - Other or unspecified |
| Asian or Asian British - Bangladeshi | Asian or Asian British - Pakistani |  | 3 | Asian or Asian British - Any other or unspecified Asian background | Asian or Asian British - South Asian |
| Asian or Asian British - Bangladeshi | Not stated |  | 38 | Asian or Asian British - Bangladeshi | Asian or Asian British - South Asian |
| Asian or Asian British - Bangladeshi | Other |  | 1 | Asian or Asian British - Bangladeshi | Asian or Asian British - South Asian |
| Asian or Asian British - Indian | Asian or Asian British - Bangladeshi |  | 6 | Asian or Asian British - Any other or unspecified Asian background | Asian or Asian British - South Asian |
| Asian or Asian British - Indian | Asian or Asian British - Pakistani |  | 19 | Asian or Asian British - Any other or unspecified Asian background | Asian or Asian British - South Asian |
| Asian or Asian British - Indian | Asian or Asian British - Sri Lankan |  | 1 | Asian or Asian British - Any other or unspecified Asian background | Asian or Asian British - South Asian |
| Asian or Asian British - Indian | Black or Black British - Any other or unspecified Black background |  | 2 | Mixed | Mixed |
| Asian or Asian British - Indian | Not stated |  | 279 | Asian or Asian British - Indian | Asian or Asian British - South Asian |
| Asian or Asian British - Indian | Other | Not stated | 1 | Asian or Asian British - Indian | Asian or Asian British - South Asian |
| Asian or Asian British - Indian | Other |  | 18 | Asian or Asian British - Indian | Asian or Asian British - South Asian |
| Asian or Asian British - Pakistani | Not stated |  | 76 | Asian or Asian British - Pakistani | Asian or Asian British - South Asian |
| Asian or Asian British - Pakistani | Other |  | 6 | Asian or Asian British - Pakistani | Asian or Asian British - South Asian |
| Asian or Asian British - Sri Lankan | Not stated |  | 1 | Asian or Asian British - Sri Lankan | Asian or Asian British - South Asian |
| Asian or Asian British - Sri Lankan | Other |  | 3 | Asian or Asian British - Sri Lankan | Asian or Asian British - South Asian |
| Black or Black British - African | Asian or Asian British - Indian |  | 4 | Mixed | Mixed |
| Black or Black British - African | Asian or Asian British - Pakistani |  | 3 | Mixed | Mixed |
| Black or Black British - African | Black or Black British - Any other or unspecified Black background | Not stated | 1 | Black or Black British - African | Black or Black British |
| Black or Black British - African | Black or Black British - Any other or unspecified Black background |  | 212 | Black or Black British - African | Black or Black British |
| Black or Black British - African | Black or Black British - Caribbean |  | 46 | Black or Black British - Any other or unspecified Black background | Black or Black British |
| Black or Black British - African | Not stated |  | 211 | Black or Black British - African | Black or Black British |
| Black or Black British - African | Other | Not stated | 2 | Black or Black British - African | Black or Black British |
| Black or Black British - African | Other |  | 32 | Black or Black British - African | Black or Black British |
| Black or Black British - Any other or unspecified Black background | Not stated |  | 35 | Black or Black British - Any other or unspecified Black background | Black or Black British |
| Black or Black British - Any other or unspecified Black background | Other |  | 8 | Black or Black British - Any other or unspecified Black background | Black or Black British |
| Black or Black British - Caribbean | Asian or Asian British - Pakistani |  | 1 | Mixed | Mixed |
| Black or Black British - Caribbean | Black or Black British - Any other or unspecified Black background | Not stated | 2 | Black or Black British - Caribbean | Black or Black British |
| Black or Black British - Caribbean | Black or Black British - Any other or unspecified Black background |  | 88 | Black or Black British - Caribbean | Black or Black British |
| Black or Black British - Caribbean | Not stated |  | 101 | Black or Black British - Caribbean | Black or Black British |
| Black or Black British - Caribbean | Other |  | 11 | Black or Black British - Caribbean | Black or Black British |
| Mixed | Asian or Asian British - Any other or unspecified Asian background | Not stated | 1 | Mixed | Mixed |
| Mixed | Asian or Asian British - Any other or unspecified Asian background |  | 49 | Mixed | Mixed |
| Mixed | Asian or Asian British - Bangladeshi |  | 2 | Mixed | Mixed |
| Mixed | Asian or Asian British - Indian |  | 22 | Mixed | Mixed |
| Mixed | Asian or Asian British - Pakistani |  | 8 | Mixed | Mixed |
| Mixed | Black or Black British - African |  | 64 | Mixed | Mixed |
| Mixed | Black or Black British - Any other or unspecified Black background |  | 13 | Mixed | Mixed |
| Mixed | Black or Black British - Caribbean |  | 38 | Mixed | Mixed |
| Mixed | Not stated |  | 160 | Mixed | Mixed |
| Mixed | Other |  | 78 | Mixed | Mixed |
| Other | Not stated |  | 160 | Other | Other |

### Trust

There were no inconsistencies across the ABS and SIP files in the trust to which each individual was assigned.

### Staff group

As indicated above, it was to be expected that some people had changed staff group over the course of the study period. However, there were 19,467 individuals with two or more different recorded values for staff group, of which 4,234 had different recorded values for staff group either in the SIP file or on the same date in the ABS file. Review of the staff group categories that occurred in combination did not identify any that clearly looked anomalous, and we therefore assumed that the individuals concerned had held two different jobs simultaneously. To account for this, we derived a new category of “Multiple” where an individual had two different staff groups on the date to which the record referred.

Where an individual was recorded as having different staff groups on different dates, each was treated as valid for the date in question, there being no basis for judging that one was implausible.

### Exposure category

As with staff group, it was to be expected that some people had changed exposure group over the course of the study period. However, there were 40,094 individuals with two or more different values for exposure category recorded, of which 7,066 were recorded simultaneously, either in the SIP file or in the ABS file. Review of the exposure codes that occurred in combination indicated that in almost all instances, one category clearly indicated greater potential for occupational exposure than the other. We therefore adopted a convention by which, as far as possible, for the date in question, the individual was assigned the higher of the two exposure categories, applying the conventions summarised in Table A6. Table A6 also indicates (in italics and underscored) combinations of exposure category for which it was less clear which was higher, and the adopted value was therefore somewhat arbitrary.

Where an individual was recorded as having different exposure categories on different dates, each was treated as valid for the date in question, there being no basis for judging that one was implausible.

**Table A6: Assigned exposure category where two jobs with different exposure categories were held simultaneously**

Assigned categories are shown in italics with an underscore where it was unclear which of a paired combination represented a higher potential for exposure to coronavirus.

Numbers in brackets indicate the frequency with which each combination of exposure categories occurred in two jobs held simultaneously (7,066 individuals had multiple exposure codes simultaneously during any time of the study of which 90 had 3 exposure codes simultaneously).

| **First exposure**  **category** | **Second exposure category** | | | | | | |
| --- | --- | --- | --- | --- | --- | --- | --- |
|  | **2** | **3** | **4** | **5** | **6** | **7** | **8** |
| **1** | 1 (207) | 1 (377) | 1 (9) | 1 (2) | 1 (80) | 1 (6) | 1 (632) |
| **2** |  | 2 (2046) | 2 (18) | 2 (7) | 2 (799) | 2 (81) | 2 (304) |
| **3** |  |  | *3* (1) | 3 (8) | 3 (663) | *3* (38) | 3 (384) |
| **4** |  |  |  | (0) | 4 (2) | (0) | 4 (2) |
| **5** |  |  |  |  | 5 (66) | *5* (5) | 5 (10) |
| **6** |  |  |  |  |  | *6* (77) | 6 (1391) |
| **7** |  |  |  |  |  |  | 7 (31) |

**Key to exposure categories:**

1. Hands-on or face-to-face care of patients much more likely to have Covid-19 than general population
2. Hands-on or face-to-face care of patients who may be more likely to have Covid-19 than general population
3. Hands-on or face-to-face care of patients whose prevalence of Covid-19 is likely to be similar to, or lower than in the general population
4. No hands-on or face-to-face care of patients, but often working in patient areas where patients are more likely to have Covid-19 than general population
5. No hands-on or face-to-face care of patients, but often working in patient areas where the prevalence of Covid-19 among patients is likely to be similar to, or lower than, in the general population
6. No hands-on or face-to-face care of patients, but occasionally in patient areas
7. Unlikely to be in patient areas, but work with material (blood/urine/clothing/equipment/installations) potentially contaminated by virus
8. Other occupation (i.e. not any of 1-7) or unknown

### Overlapping spells of absence in the same individual

There were 63,556 instances in which two or more spells of absence in the same individual overlapped. For 28,217 recordings, the same absence was recorded for different jobs held simultaneously. Among the remainder, in most cases (n = 29,835), one was nested within the other, but in 5,504 a second absence started during the first, and ended after it or had a missing end date. Review of the reasons ascribed to these overlapping absences indicated that they were generally plausible. Sickness absences cannot overlap with other sickness absences, only non-sickness absences can overlap with sickness absences or non-sickness absence, and we only observed the later (e.g. an episode of absence for infection precaution or medical suspension superimposed on a spell of absence for another reason). However, there was only one instance of overlap between spells of sickness absence for two different specific disease categories.

We therefore made no changes to the records in the ABS file in response to overlapping episodes of absence. However, we decided that in subsequent analyses:

a) When calculating period of absence from work irrespective of reason (e.g. to exclude people who were absent continuously throughout a specified period), we would treat overlapping periods of absence as a single episode running from the earliest start date to latest end date for the overlapping episodes.
b) Where the focus was on spells of absence for a specific reason (e.g. sickness absence for Covid-19), we would ignore overlaps with periods of absence for other reasons, and when multiple periods of absence for the reason of interest overlapped, we would treat them as a single episode of absence for the reason of interest, running from the earliest start date to the latest end date of the overlapping spells.
