## Supplementary File B1 for "Occupational risks of COVID-19 in NHS workers in England"

Supplementary File B

SPECIFICATION OF STUDY PERIOD, COVID-19 SICKNESS ABSENCE AND OCCUPATIONAL VARIABLES

### Study period

National data on hospital admissions for Covid-19 by trust^B1^ indicated that they first began about 19 March, and because of the lag between onset of illness and admission to hospital, it was to be expected that infection first started to emerge on any scale a week or two earlier. Furthermore, analysis of start dates for Covid-19 sickness absence (for definition see below) indicated a marked increase in daily numbers from 9 March. Also, analysis of results from antibody tests during late May to July in the subset of cohort members at Guys and St Thomas’s Trust, showed that the ratio of positive to negative tests among people whose only episode of Covid-19 sickness absence began during the seven days from 9 March was substantially higher than that in people who had taken no such absence at any time.

The start of the study period was therefore taken as 9 March 2020. The end date of 31 July 2020 was that up to which complete data were available in the databases from the NHS Electronic Staff Record.

### Covid-19 sickness absence

We sought measures of sickness absence that would serve as markers, albeit imperfect, for incident Covid-19 and for incident cases of more severe Covid-19. As well as the three variables that were used to specify “Collapsed absence category”, the ABS file included a variable that denoted whether absences were related to Covid-19. This had five possible values: No relation recorded; Coronavirus (COVID-19); Coronavirus (COVID-19) – household member symptoms; Coronavirus (COVID-19) – post travel quarantine; and Coronavirus (COVID-19) – test and trace contact. The last three categories seemed unlikely to reflect illness in the staff member. However, it was possible that they were not used consistently, and that some absences to which they could have been applied, were instead assigned to the less specific category labelled simply as Coronavirus (COVID-19).

Among the 102,422 sickness absence episodes that began during the study period, and were labelled as Coronavirus (COVID-19), the large majority fell in the collapsed absence categories “Sickness – chest and respiratory” (n = 33,555, mean duration 13 days, median duration 8 days), “Sickness – cough, flu” (n = 27,937, mean duration 12 days, median duration 7 days) and “Sickness – infectious disease” (n= 39,110, mean duration 13 days, median duration 8 days). The similarity of the mean and median durations across the three categories suggested that they had been applied to similar types of illness. In addition, there were two less specific collapsed absence categories, which when associated with a label of Coronavirus (COVID-19), had comparable mean and median durations. These were “Sickness – other” (n= 430, mean duration 15 days, median duration 9 days), and “Sickness – unknown” (n= 668, mean duration 14 days, median duration 8 days). Together, these five categories were therefore aggregated as “Covid-19 sickness absence”.

The label, Coronavirus (COVID-19) also occurred in association with other categories of sickness absence, but these were less frequent, and had different durations (e.g. “Sickness – mental health”, n = 158, mean duration 52 days, median duration 42.5 days, “Sickness – part day”, n = 69, mean duration 1 day, median duration 1 day). It was less certain that such absences indicated an episode of illness from Covid-19, and we therefore excluded them from our definition of “Covid-19 sickness absence”.

Table B1 summarises the durations of the 101,700 episodes of Covid-19 sickness absence which started during the study period. Based on this information, we classed episodes of Covid-19 sickness absence as “prolonged” if their duration exceeded 14 days.

### Table B1. Durations of Covid-19 sickness absence

|  | N | % | Minimum | 1st Quartile | Median | Mean | 3rd Quartile | Maximum |
| --- | --- | --- | --- | --- | --- | --- | --- | --- |
| All | 101700 | 100 | 1 | 6 | 8 | 12.41 | 14 | 144 |
| ≤7 days | 50664 | 49.8 | 1 | 3 | 6 | 5.19 | 7 | 7 |
| 8-14 days | 29414 | 28.9 | 8 | 9 | 11 | 11.02 | 14 | 14 |
| 15-21 days | 10645 | 10.5 | 15 | 15 | 17 | 17.41 | 19 | 21 |
| >21 days | 10977 | 10.8 | 22 | 26 | 33 | 44.78 | 52 | 144 |

### Staff group and exposure category

Both of these variables could vary over time (see Supplementary File A). In the analysis for this paper, we aimed to classify individuals according to the job that they held on 9 March 2020. Where staff group and/or exposure category changed over time, we therefore gave preference in the following order:

- That from the absence record with the most recent start date at 9 March 2020
- If there was no absence starting before 9 March 2020, that from the absence record with the earliest start date after 9 March 2020
- If there were no absences at all, that from the SIP file

### References

B1.	COVID-19 NHS Situation Reports, COVID-19 daily situation report. Data as reported on 05-Nov-20. Published: 12 November 2020. Downloaded from: <https://www.england.nhs.uk/statistics/statistical-work-areas/covid-19-hospital-activity/>
