## Supplementary File C1 for "Occupational risks of COVID-19 in NHS workers in England"

Supplementary File C

SENSITIVITY ANALYSES excluding individuals WITH imputed or missing data, or multiple jobs

### Table C1. Associations of risk factors at baseline with start of an episode of Covid-19 sickness absence (any and prolonged) during 9 March to 31 July 2020 - excluding individuals with imputed or missing data, or multiple jobs

Excludes 6,554 individuals for whom one or more of age, sex or ethnicity was imputed because of inconsistencies in the raw data, 3 with unknown staff group, 44,134 with multiple staff group or exposure categories during the study period (either because they held multiple jobs simultaneously or changed their job), and 1,923 with a missing or imputed end date of an absence.

Risk estimates were derived from two logistic regression models that included all of the variables for which results are presented, together with trust (191 categories), and are relative to no Covid-19 sickness absence. An episode of Covid-19 sickness absence was classed as prolonged if it had lasted >14 days by 31 July 2020.

|  | **Any Covid-19 sickness absence** | | | |  | **Any prolonged Covid-19 sickness absence** | | | |
| --- | --- | --- | --- | --- | --- | --- | --- | --- | --- |
| **Risk factor** | **Model 1** | | **Model 2** | |  | **Model 1** | | **Model 2** | |
|  | **RR** | **(95% CI)** | **RR** | **(95% CI)** |  | **RR** | **(95% CI)** | **RR** | **(95% CI)** |
| **Sex** |  |  |  |  |  |  |  |  |  |
| Female | *ref.* | *ref.* | *ref.* | *ref.* |  | *ref.* | *ref.* | *ref.* | *ref.* |
| Male | 1.01 | 0.99 - 1.03 | 1.02 | 1.00 - 1.04 |  | 1.02 | 0.98 - 1.06 | 1.03 | 0.99 - 1.08 |
| **Age (years)** |  |  |  |  |  |  |  |  |  |
| <30 | *ref.* | *ref.* | *ref.* | *ref.* |  | *ref.* | *ref.* | *ref.* | *ref.* |
| 30-34 | 0.97 | 0.94 - 1.00 | 0.97 | 0.94 - 1.00 |  | 1.26 | 1.17 - 1.36 | 1.26 | 1.17 - 1.35 |
| 35-39 | 0.98 | 0.95 - 1.01 | 0.99 | 0.96 - 1.02 |  | 1.55 | 1.44 - 1.66 | 1.55 | 1.44 - 1.67 |
| 40-44 | 0.99 | 0.97 - 1.02 | 1.00 | 0.97 - 1.03 |  | 1.72 | 1.61 - 1.84 | 1.71 | 1.60 - 1.84 |
| 45-49 | 1.00 | 0.98 - 1.03 | 1.01 | 0.98 - 1.04 |  | 2.01 | 1.88 - 2.15 | 1.99 | 1.86 - 2.12 |
| 50-54 | 0.97 | 0.95 - 1.00 | 0.98 | 0.95 - 1.01 |  | 2.15 | 2.01 - 2.29 | 2.13 | 2.00 - 2.28 |
| 55-60 | 0.88 | 0.86 - 0.91 | 0.88 | 0.86 - 0.91 |  | 2.07 | 1.94 - 2.21 | 2.05 | 1.92 - 2.20 |
| >60 | 0.75 | 0.72 - 0.78 | 0.74 | 0.72 - 0.77 |  | 2.10 | 1.95 - 2.26 | 2.06 | 1.91 - 2.22 |
| **Ethnicity** |  |  |  |  |  |  |  |  |  |
| White | *ref.* | *ref.* | *ref.* | *ref.* |  | *ref.* | *ref.* | *ref.* | *ref.* |
| South Asian | 1.42 | 1.38 - 1.46 | 1.40 | 1.36 - 1.44 |  | 2.56 | 2.43 - 2.70 | 2.49 | 2.36 - 2.62 |
| Other or unspecified Asian | 1.72 | 1.66 - 1.77 | 1.64 | 1.59 - 1.69 |  | 2.93 | 2.78 - 3.10 | 2.71 | 2.56 - 2.86 |
| Black | 1.15 | 1.11 - 1.18 | 1.13 | 1.10 - 1.17 |  | 1.73 | 1.64 - 1.84 | 1.69 | 1.60 - 1.79 |
| Mixed | 1.14 | 1.07 - 1.20 | 1.13 | 1.07 - 1.20 |  | 1.39 | 1.24 - 1.56 | 1.37 | 1.22 - 1.54 |
| Other | 1.48 | 1.40 - 1.56 | 1.43 | 1.36 - 1.51 |  | 2.42 | 2.21 - 2.65 | 2.29 | 2.09 - 2.51 |
| Unknown | 1.07 | 1.03 - 1.11 | 1.07 | 1.03 - 1.11 |  | 1.31 | 1.21 - 1.42 | 1.31 | 1.21 - 1.42 |
| **Episodes of sickness absence in 2019** |  |  |  |  |  |  |  |  |  |
| 0 | *ref.* | *ref.* | *ref.* | *ref.* |  | *ref.* | *ref.* | *ref.* | *ref.* |
| 1 | 1.39 | 1.36 - 1.42 | 1.38 | 1.35 - 1.41 |  | 1.49 | 1.43 - 1.56 | 1.48 | 1.42 - 1.55 |
| 2-3 | 1.82 | 1.78 - 1.86 | 1.80 | 1.76 - 1.83 |  | 2.00 | 1.92 - 2.08 | 1.97 | 1.89 - 2.05 |
| >3 | 2.41 | 2.35 - 2.47 | 2.37 | 2.32 - 2.43 |  | 2.63 | 2.50 - 2.77 | 2.57 | 2.44 - 2.70 |
| **Staff group at 9 March 2020** |  |  |  |  |  |  |  |  |  |
| Administrative and clerical | *ref.* | *ref.* | *ref.* | *ref.* |  | *ref.* | *ref.* | *ref.* | *ref.* |
| Additional clinical services | 2.30 | 2.24 - 2.36 | 1.62 | 1.54 - 1.71 |  | 2.93 | 2.78 - 3.09 | 1.82 | 1.61 - 2.04 |
| Additional professional scientific and technical | 1.36 | 1.31 - 1.43 | 1.04 | 0.98 - 1.12 |  | 1.19 | 1.07 - 1.33 | 0.97 | 0.83 - 1.13 |
| Allied health professionals | 1.95 | 1.89 - 2.02 | 1.33 | 1.26 - 1.41 |  | 1.73 | 1.60 - 1.87 | 1.06 | 0.93 - 1.21 |
| Estates and ancillary | 1.44 | 1.39 - 1.50 | 1.29 | 1.24 - 1.34 |  | 1.60 | 1.48 - 1.73 | 1.39 | 1.28 - 1.51 |
| Healthcare scientists | 1.17 | 1.10 - 1.24 | 1.03 | 0.95 - 1.11 |  | 0.96 | 0.83 - 1.11 | 0.88 | 0.73 - 1.05 |
| Medical and dental | 1.54 | 1.49 - 1.60 | 1.08 | 1.01 - 1.14 |  | 1.05 | 0.97 - 1.15 | 0.69 | 0.60 - 0.79 |
| Nursing and midwifery registered | 2.28 | 2.23 - 2.33 | 1.57 | 1.49 - 1.65 |  | 2.63 | 2.49 - 2.76 | 1.54 | 1.37 - 1.73 |
| Students | 1.88 | 1.60 - 2.20 | 1.34 | 1.13 - 1.59 |  | 1.73 | 1.03 - 2.91 | 1.19 | 0.70 - 2.02 |
| **Exposure category at 9 March 2020*** |  |  |  |  |  |  |  |  |  |
| Care of patients much more likely to have  Covid-19 than general population | - | - | 1.49 | 1.40 - 1.58 |  | - | - | 1.54 | 1.34 - 1.77 |
| Care for patients who may be more likely  to have Covid-19 than general population | - | - | 1.42 | 1.35 - 1.50 |  | - | - | 1.71 | 1.52 - 1.93 |
| Care of patients with similar or lower prevalence  of Covid-19 than general population | - | - | 1.06 | 1.00 - 1.12 |  | - | - | 1.05 | 0.93 - 1.18 |
| No patient care but often in areas where  patients have higher prevalence of Covid-19  than general population | - | - | 0.75 | 0.54 - 1.05 |  | - | - | 1.12 | 0.57 - 2.19 |
| No patient care but often in areas where  patients have similar or lower prevalence of  Covid-19 than general population | - | - | 1.29 | 1.19 - 1.39 |  | - | - | 0.97 | 0.80 - 1.17 |
| No patient care, occasionally in patient areas | - | - | *ref.* | *ref.* |  | - | - | *ref.* | *ref.* |
| Unlikely to be in patient areas, but work with  material potentially contaminated by coronavirus | - | - | 0.92 | 0.85 - 0.99 |  |  |  | 0.79 | 0.67 - 0.94 |
| Other or unknown | - | - | 0.73 | 0.70 - 0.76 |  |  |  | 0.66 | 0.60 - 0.72 |

*Exposure categories are based on the constructed Job Exposure Matrix (JEM). For more information see Supplementary file A.
